# Machine-learning analysis of temporal molecular dynamics stratifies autism likelihood - a multinational study

**DOI:** 10.1101/2025.11.19.25340581

**Authors:** Vishal Midya, Ghalib A. Bello, Louis A. Gomez, Manuel Ruiz Marin, Sujeewa C. Piyankarage, Suzy Elhlou, Jyoti Chumber, Juliet Jaramilo, Sophie Dessalle, Maayan Yitshak-Sade, Avi Reichenberg, Alejandra Cantoral, Rosalind J. Wright, Robert Wright, Shoji Nakayama, Deborah H. Bennett, Rebecca J. Schmidt, Sven Bölte, Manish Arora

## Abstract

Absence of autism risk-stratification tools under 18 months hampers early intervention. In a multinational sample of 1697 participants, aged one month and older, we provide proof-of-concept that temporal molecular dynamics can stratify autism likelihood. Using laser-ablation-inductively-coupled-plasma-mass-spectrometry, we measured elemental intensities along growth increments of single hair strands at ∼800 timepoints. We developed a first-stage model to stratify individuals into a lower autism probability group and applied a second-stage model to the remaining participants, stratifying them into intermediate- and high-probability groups. Models were trained, ensembled, and tuned on participants from California and Sweden, then tested on 580 participants (within- and external-population replication in New York, Mexico, and Japan). Likelihood ratios (95%CI) for autism in low-, intermediate-, and high-probability groups were 0.18(0.15-0.23), 1.09(0.99-1.20), and 2.62(1.55-4.00), respectively. Low-probability classification (first-stage) had sensitivity of 96%(0.91-0.98), and high-probability classification (second-stage) had specificity of 90%(0.86-0.92). Our data support that elemental biodynamics can objectively stratify autism likelihood.

---

Autism is now diagnosed in approximately 1 to 3% of all children in most mid to high-income countries.^1,2^ Due to increasing referral rates and persistent shortages in clinicians specializing in autism diagnosis, wait times for autism diagnostic assessment may be months to years.^3,4^ As a result, the median age of diagnosis in the US and other nations remains over 4 years, hindering timely intervention. Environmental factors have been shown to moderate or mediate neurodevelopment and autism phenotypes both directly and indirectly through gene-environment interactions.^5–7^ Furthermore, diagnosis rates of autism in the US and elsewhere in the world are on the rise (recent data showing 1 in 31 children are diagnosed with autism in the US), and multiple other factors besides genetics, such as environmental exposures, improved screening and diagnostic practices, are likely at play.

Among those environmental factors, exposure to toxic metals and deficiencies in nutritional elements during prenatal and early childhood have been associated with increased likelihood of autism and other developmental conditions.^8,9^ Animal studies suggest that elemental exposure can alter brain development by influencing neurotransmission in frontal and subcortical brain regions, some of which are also implicated in autism.^8,10^ Elemental biodynamics in humans, i.e., the dynamic processes by which homeostasis of essential and non-essential elements is maintained within the body’s systems, are tightly regulated.^11–13^ These processes are controlled by intricate regulatory mechanisms that respond to various environmental and internal signals, ensuring that the concentrations of these elements remain within a physiological range.^14,15^ Key biological processes (metabolism, acid-base balance, renal function, for example) cyclically regulate the temporal dynamics of elemental stoichiometry to maintain homeostasis.^16–18^ Some of these processes are also implicated in changes of amino acid pathways and mitochondrial alterations, and can contribute to autism-related behaviors.^19,20^ One hypothesized mechanism underlying divergent neurotypes, therefore, involves elemental homeostasis. The biodynamics of elemental homeostasis guiding early development are tightly controlled by regulatory mechanisms that respond to external environmental perturbations.^21,22^ Therefore, biodynamic synchronization in elemental profiles may serve as markers of homeostatic regulation that may be predictive of present and future likelihood of autism-related behaviors.^23^

Fluid-based biomarkers (e.g., blood and urine) provide a “snapshot” of a given time point. Obtaining temporal intensity patterns by collecting repeated blood, urine, or stool samples is generally not feasible due to safety and logistical barriers. To overcome this challenge, we analyzed elemental intensities in hair growth increments using laser-ablation-inductively coupled plasma-mass-spectrometry (LA-ICP-MS) to generate time-resolved signals of elemental patterns at approximately hourly resolution along the hair shaft, with an average of ∼800 sampling points per hair strand.^24^ Since hair grows at ∼1 cm a month, these data represent elemental concentration patterns over time. Using a machine learning (ML) architecture, we leveraged this time-series data to study elemental biodynamics as a biomarker of homeostasis that can distinguish individuals diagnosed with autism from those without. In this study, we develop and validate a novel diagnostic aid that leverages the temporal biodynamics of 12 elements: magnesium (Mg), phosphorus (P), calcium (Ca), copper (Cu), zinc (Zn), strontium (Sr), barium (Ba), lead (Pb), lithium (Li), manganese (Mn), Sulphur (S) and Arsenic (As).

When considering children referred from general pediatricians for autism evaluation by specialists, a significant proportion of children do not ultimately get diagnosed with autism, but all children undergo lengthy behavioral assessments that place demands on clinician time and burden on service users.^25,26^ Therefore, there is a need for objective tools that stratify autism likelihood and create a sequential, objective probability-based priority score to lessen the burden on clinicians’ time. Here, using non-invasively collected hair strands, we devise and describe a novel machine-learning-driven two-stage triage approach that exploits the time-varying biodynamics of elemental intensities to stratify autism likelihood. Figure 1 illustrates the overall conceptual framework for this study, as well as the overview of the results for autism likelihood stratification.

**Figure 1:**
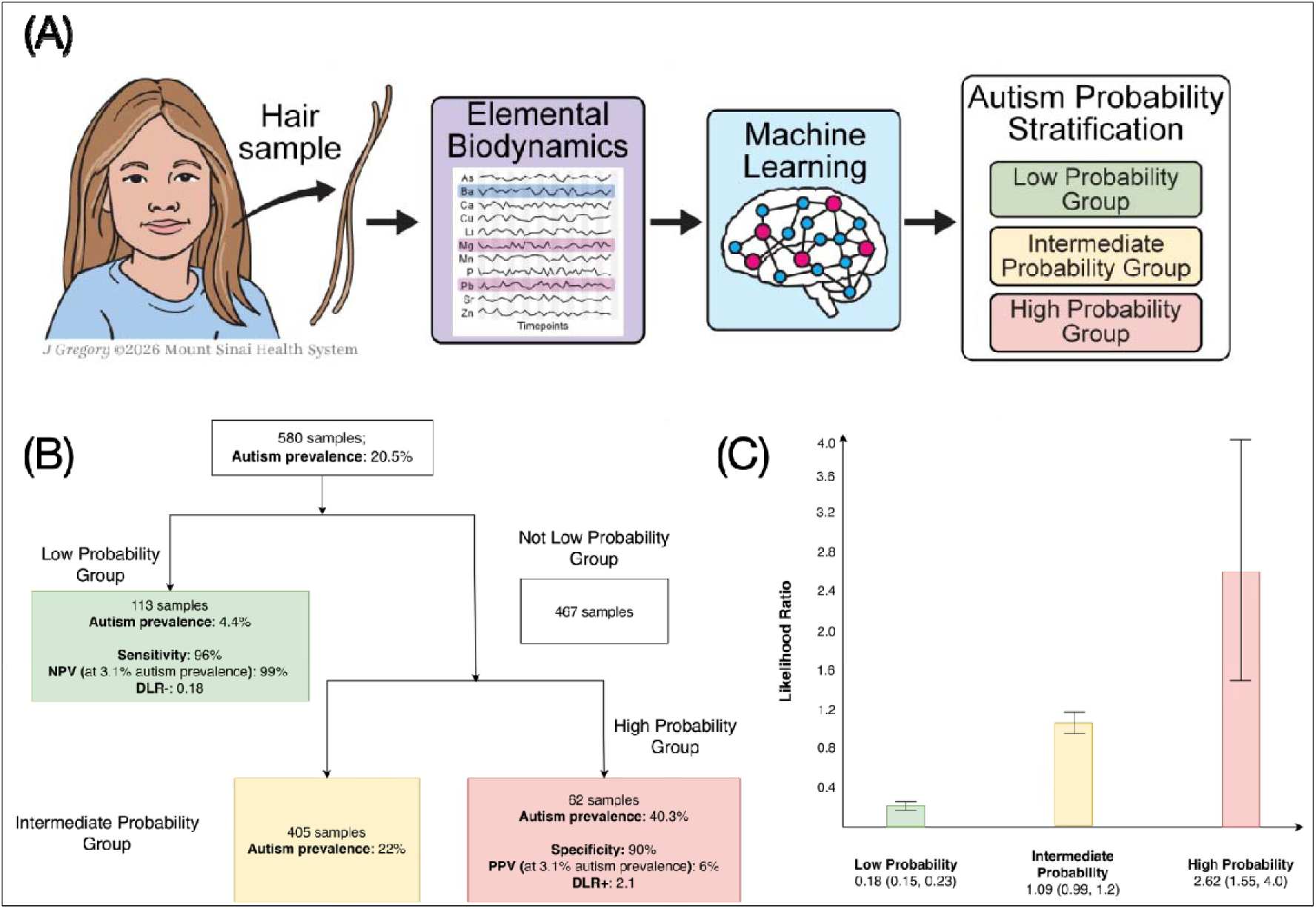
Overall schematics, flow, and results of hair-based elemental biodynamics model. (A) illustrates the conceptual design of the stratified autism likelihood groups. (B-C) Summary of the two-stage stratified triage system in the validation samples (n = 580) showing (B) high-level performance metrics at each stage of stratification (i.e., low, intermediate, and high probability groups), and (C) bar plots of group-specific likelihood ratios (with their 95% CI) for each group. In both figures, green, yellow, and red denote low-, medium-, and high-autism-likelihood groups, respectively. PPV: Positive predictive value; NPV: Negative predictive value; DLR-: Diagnostic likelihood ratio (positive); DLR-: Diagnostic likelihood ratio (negative).

## RESULTS

To test our hypothesis, we studied a multinational sample of children (N=1697) from diverse populations across multiple countries, including the United States, Sweden, Mexico, and Japan encompassing six cohorts: CHARGE (n = 825) and MARBLES (n = 287) from California, RATSS from Sweden (n = 306), Seaver (n = 39) and PRISM (n = 84) cohorts from New York, JECS from Japan (n = 110), and a cohort from Mexico (n = 46). Among 1697 samples, 354 were autism cases, and 1343 were non-autism controls. Only one hair strand was used per participant (see Table 3 in the Methods). All autism assessments followed DSM-5 criteria supported by standard diagnostic instruments (ADOS-2 and/or ADI-R). Figure 2 shows an overall schematic of how different cohorts were used for training, stacking, tuning, and validation in the two-stage models (see Methods for more details).

**Figure 2:**
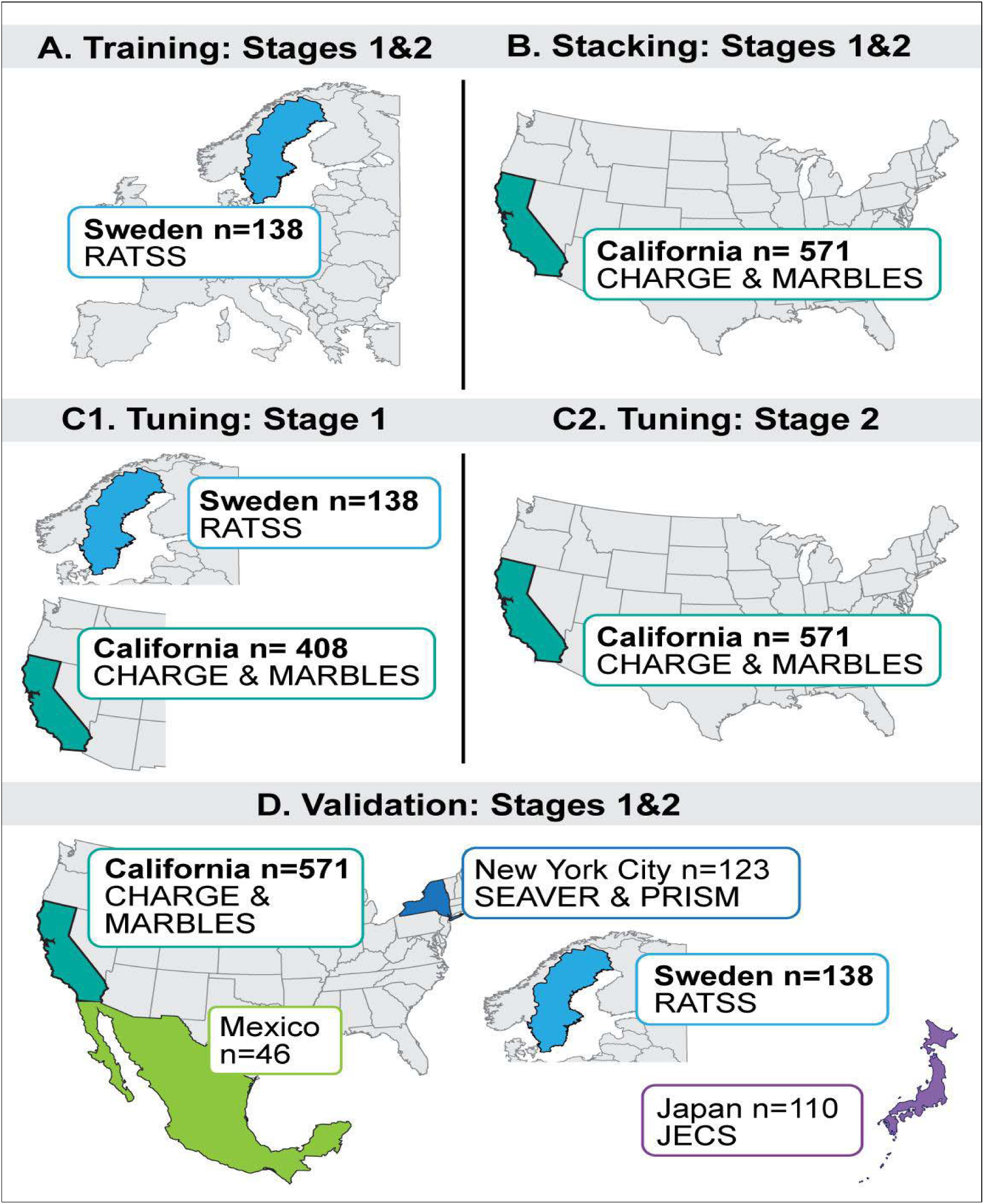
Overall schematic of cohorts used in training, stacking, tuning, and validation tasks. In training, data from the RATSS cohort (n = 138, Sweden) were used for both Stage 1 and 2 models. During the stacking phase, data from the CHARGE and MARBLES cohort (n = 571, California) were used for both Stage 1 and 2 models. For the Stage 1 model, the threshold was tuned using the same RATSS cohort (n = 138) used for training, as well as additional data from the CHARGE and MARBLES cohorts (n = 408, California) that included a large number of controls. The increased proportion of controls was used to tune threshold for the left tail of the Stage 1 model, which was intended for the low-probability group. For the Stage 2 model, the threshold was tuned using data from the CHARGE and MARBLES cohorts (n = 571, California), which were previously used for stacking. Lastly, both the Stage 1 and 2 models were sequentially tested on additional unseen CHARGE and MARBLES (California), and RATSS (Sweden) data, as well as external validation data from Mexico City (n = 46), New York City (n = 123), and Japan (n = 110).

**Table 1:**
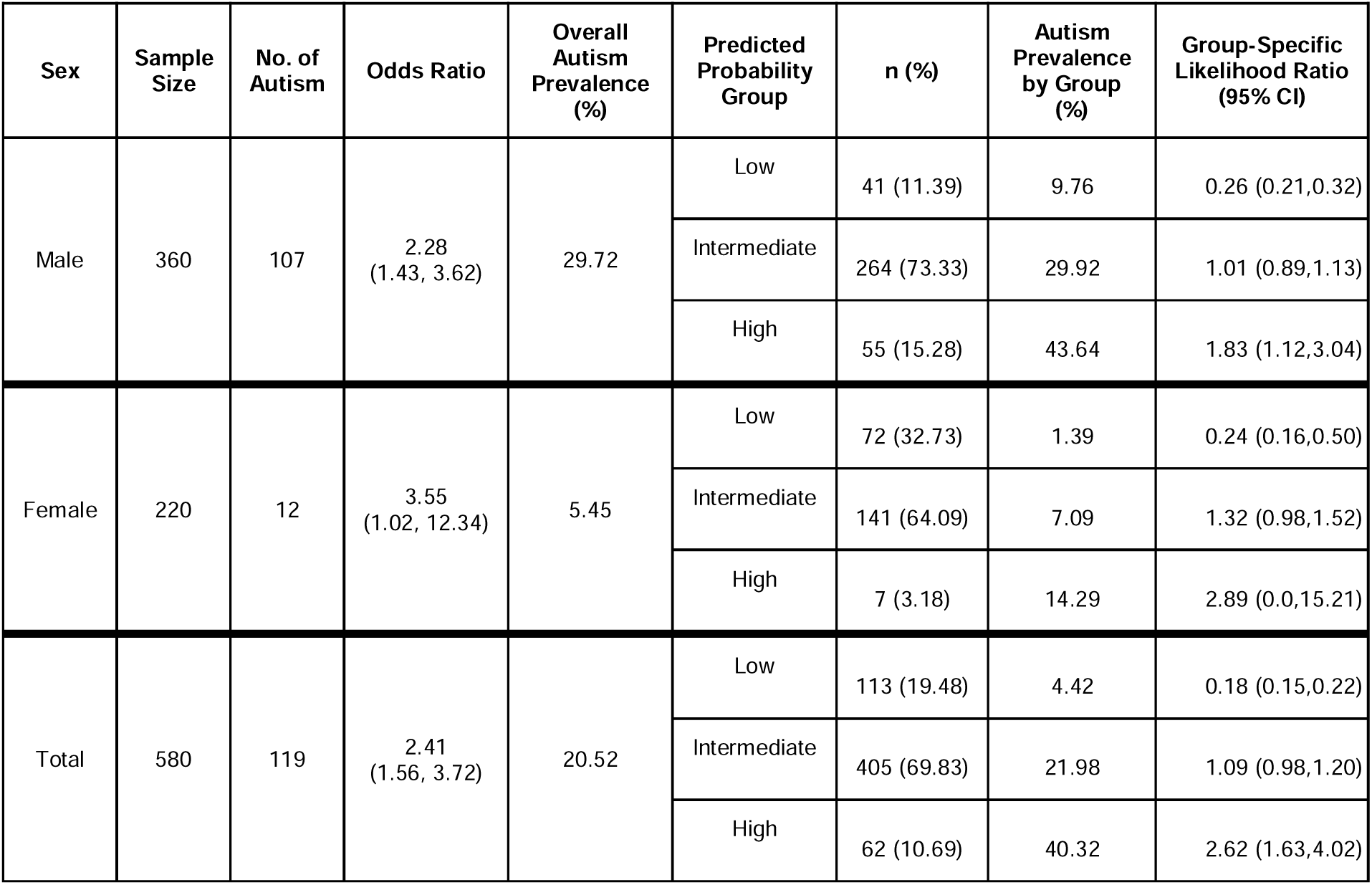
Overall performance metrics for the likelihood stratification framework.

| Sex | Sample Size | No. of Autism | Odds Ratio | Overall Autism Prevalence (%) | Predicted Probability Group | n (%) | Autism Prevalence by Group (%) | Group-Specific Likelihood Ratio (95% CI) |
| --- | --- | --- | --- | --- | --- | --- | --- | --- |
| Male | 360 | 107 | 2.28<br>(1.43, 3.62) | 29.72 | Low | 41 (11.39) | 9.76 | 0.26 (0.21,0.32) |
|  |  |  |  |  | Intermediate | 264 (73.33) | 29.92 | 1.01 (0.89,1.13) |
|  |  |  |  |  | High | 55 (15.28) | 43.64 | 1.83 (1.12,3.04) |
| Female | 220 | 12 | 3.55<br>(1.02, 12.34) | 5.45 | Low | 72 (32.73) | 1.39 | 0.24 (0.16,0.50) |
|  |  |  |  |  | Intermediate | 141 (64.09) | 7.09 | 1.32 (0.98,1.52) |
|  |  |  |  |  | High | 7 (3.18) | 14.29 | 2.89 (0.0,15.21) |
| Total | 580 | 119 | 2.41<br>(1.56, 3.72) | 20.52 | Low | 113 (19.48) | 4.42 | 0.18 (0.15,0.22) |
|  |  |  |  |  | Intermediate | 405 (69.83) | 21.98 | 1.09 (0.98,1.20) |
|  |  |  |  |  | High | 62 (10.69) | 40.32 | 2.62 (1.63,4.02) |

**Table 2:** Stage-wise metrics for our autism likelihood stratification framework.

| Stage 1: Low-Likelihood Stratification | Sex | Sample Size | No. of Autism (% autism) | Sensitivity (95% CI) | DLR- |
| --- | --- | --- | --- | --- | --- |
|  | Male | 360 | 107 (29.72) | 0.96 (0.91, 0.99) | 0.26 |
|  | Female | 220 | 12 (5.45) | 0.92 (0.65, 0.99) | 0.24 |
|  | Total | 580 | 119 (20.52) | 0.96 (0.91, 0.98) | 0.18 |
| Stage 2: High- and Intermediate-Likelihood Stratification | Sex | Sample Size | No. of Autism | Specificity (95% CI) | DLR+ |
|  | Male | 319 | 103 (32.29) | 0.86 (0.81,0.90) | 1.62 |
|  | Female | 148 | 11 (7.43) | 0.96 (0.92,0.99) | 2.08 |
|  | Total | 467 | 114 (24.41) | 0.90 (0.86,0.92) | 2.09 |

DLR+: Diagnostic likelihood ratio (positive); DLR-: Diagnostic likelihood ratio (negative).

**Table 3:** Demographic and elemental statistics for all samples included in the study.

|  | All<br>(n=1697) | Autism<br>(n=354) | Non-autism<br>(n=1343) | p-value* |
| --- | --- | --- | --- | --- |
| **CHARGE (California, US, n=825, Autism=164, non-autism=661)** |  |  |  |  |
| **Age (in months) at hair collection, median (IQR)** | 36 (20) | 27 (20) | 41 (19) | <0.01 |
| **Sex, n (%)** |  |  |  | <0.01 |
| Female | 189 (22.91%) | 17 (10.37%) | 172 (26.02%) |  |
| Male | 636 (77.09%) | 147 (89.63%) | 489 (73.98%) |  |
| **Hair length (in centimeters), Mean (SD)** | 1.19 (0.01) | 1.19 (0.01) | 1.19 (0.01) | 0.12 |
| **Hair length (in days of growth), Mean (SD)** | 35.72 (0.43) | 35.76 (0.39) | 35.71 (0.44) | 0.12 |

| **MARBLES (California, US, n=287, Autism=66, non-autism=221)** |  |  |  |  |
| --- | --- | --- | --- | --- |
| **Age (in months) at hair collection, median (IQR)** | 12.5 (11.7) | 12.7 (12.0) | 12.5 (11.1) | 0.23 |
| **Sex, n (%)** |  |  |  | < 0.01 |
| Female | 127 (44.3%) | 17 (25.7%) | 110 (49.8%) |  |
| Male | 160 (55.8%) | 49 (74.2%) | 111 (50.2%) |  |
| **Hair length (in centimeters), Mean (SD)** | 1.19 (0.02) | 1.19 (0.02) | 1.19 (0.02) | 0.36 |
| **Hair length (in days of growth), Mean (SD)** | 35.64 (0.47) | 35.65 (0.49) | 35.64 (0.47) | 0.36 |
| **RATSS (Sweden, n=306, Autism=70, non-autism=236)** |  |  |  |  |
| **Age (in months) at hair collection in months, median (IQR)** | 168 (84) | 156.0 (48) | 186.0 (84) | <0.01 |
| **Sex, n (%)** |  |  |  | <0.01 |
| Female | 136 (44.44%) | 21 (30.00%) | 115 (48.73%) |  |
| Male | 170 (55.56%) | 49 (70.00%) | 121 (51.27%) |  |
| **Hair length (in centimeters), Mean (SD)** | 0.91 (0.29) | 0.80 (0.31) | 0.94 (0.28) | <0.01 |
| **Hair length (in days of growth), Mean (SD)** | 27.16 (8.84) | 24.13 (9.22) | 28.07 (8.54) | <0.01 |
| **SEAVER (New York City, US, n=39, Autism=21, non-autism=18)** |  |  |  |  |
| **Age (in months) at hair collection, median (IQR)** | 72 (54) | 72 (72) | 90 (48) | 0.81 |
| **Sex, n (%)** |  |  |  | <0.01 |
| Female | 14 (35.90%) | 3 (14.29%) | 11 (61.11%) |  |
| Male | 25 (64.10%) | 18 (85.71%) | 7 (38.89%) |  |
| **Hair length (in centimeters), Mean (SD)** | 1.19 (0.01) | 1.19 (0.00) | 1.19 (0.01) | 0.38 |
| **Hair length (in days of growth), Mean (SD)** | 35.70 (0.17) | 35.70 (0.07) | 35.71 (0.24) | 0.38 |
| **Mexico (Mexico City, n=46, Autism=33, non-autism=13)** |  |  |  |  |
| **Age (in months) at hair collection, median (IQR)** | 35.5 (22.75) | 38.0 (25.0) | 30.0 (19.0) | 0.03 |
| **Sex, n (%)** |  |  |  | <0.01 |
| Female | 12 (26.09%) | 4 (12.12%) | 8 (61.54%) |  |
| Male | 34 (73.91%) | 29 (87.88%) | 5 (38.46%) |  |
| **Hair length (in centimeters), Mean (SD)** | 1.16 (0.02) | 1.17 (0.01) | 1.15 (0.02) | 0.01 |
| **Hair length (in days of growth), Mean (SD)** | 34.82 (0.58) | 34.98 (0.45) | 34.40 (0.66) | 0.01 |
| **PRISM (New York City, US, n=84, Autism=0, non-autism=84)** |  |  |  |  |
| **Age (in months) at hair collection, median (IQR)** | 48.0 (7.25) | - | 48.0 (7.25) | - |
| **Sex, n (%)** |  |  |  |  |
| Female | 33 (39.29%) | - | 33 (39.29%) | - |
| Male | 51 (60.71%) | - | 51 (60.71%) | - |
| **Hair length (in centimeters), Mean (SD)** | 1.16 (0.01) | - | 1.16 (0.01) | - |
| **Hair length (in days of growth), Mean (SD)** | 34.75 (2.87) | - | 34.75 (2.87) | - |
| **JECS (Japan, n=110, Autism=0, non-autism=110)** |  |  |  |  |
| **Age (in months) at hair collection, median (IQR)** | 1.0 (0.0) |  | 1.0 (0.0) | - |
| **Sex, n (%)** |  |  |  |  |
| Female | 55 (50%) |  | 55 (50%) |  |
| Male | 55 (50%) |  | 55 (50%) |  |
| **Hair length (in centimeters), Mean (SD)** | 1.14 (0.11) | - | 1.14 (0.11) | - |
| **Hair length (in days of growth), Mean (SD)** | 34.16 (3.20) | - | 34.16 (3.20) | - |

\*The p-value was calculated using the chi-squared test for categorical variables (except in cases of low cell counts, in which case Fisher's Exact test was used) and the Mann-Whitney U test for continuous variables.

### Elemental Biodynamics

The time-varying elemental intensity of 12 elements [magnesium (Mg), phosphorus (P), calcium (Ca), copper (Cu), zinc (Zn), strontium (Sr), barium (Ba), lead (Pb), lithium (Li), manganese (Mn), Sulphur (S), and Arsenic (As)] were used to construct features that capture the essence of dynamic changes in intensity levels. Along with considering temporally independent features (e.g., measures of the central tendencies of intensity distributions), we estimated several features that capture the dynamic essence of temporality. The current literature shows that measuring elemental intensities over time in hair strands may serve as a biomarker of altered elemental homeostasis.^24^ Since elemental homeostasis is tightly controlled by stable regulatory mechanisms that respond to environmental signals, we hypothesized that the longitudinal dynamic patterns of elemental intensities (henceforth referred to as elemental biodynamics) may serve as a marker of homeostatic alterations.^12^ Therefore, delineating such measures of elemental homeostasis may serve as a predictor of disorders.

To capture diverse aspects of temporal biodynamics measured using our hair biomarkers, we computed multiple feature-extraction strategies, including nonlinear recurrence-structure (RQA), cross-element synchronization (CRQA and multiscale cross-entropy), entropy and complexity measures, distributional descriptors (e.g., summary statistics), and network-derived centrality measures.^27–30^ These approaches quantified various properties of the elemental time series, such as rhythmicity, irregularity, cross-element synchronicity, and statistical properties. See supplementary section S3 for full details on these feature engineering methods. To illustrate how inter-element relationships and higher-order interaction patterns differ between autism cases and controls, and by participant sex, we present a specific feature type (elemental network centrality) to visualize elemental interaction networks from validation samples (see Figure 3). Note that compared to non-autism controls, the autism cases have two separate connections among elements. Although presented only as an illustration, our modelling strategy incorporates such nuanced differences in identifying features that can differentiate autism cases from controls. These examples highlight variability in network topology and demonstrate the type of inter-element interactions encoded within our modeling framework.

**Figure 3:**
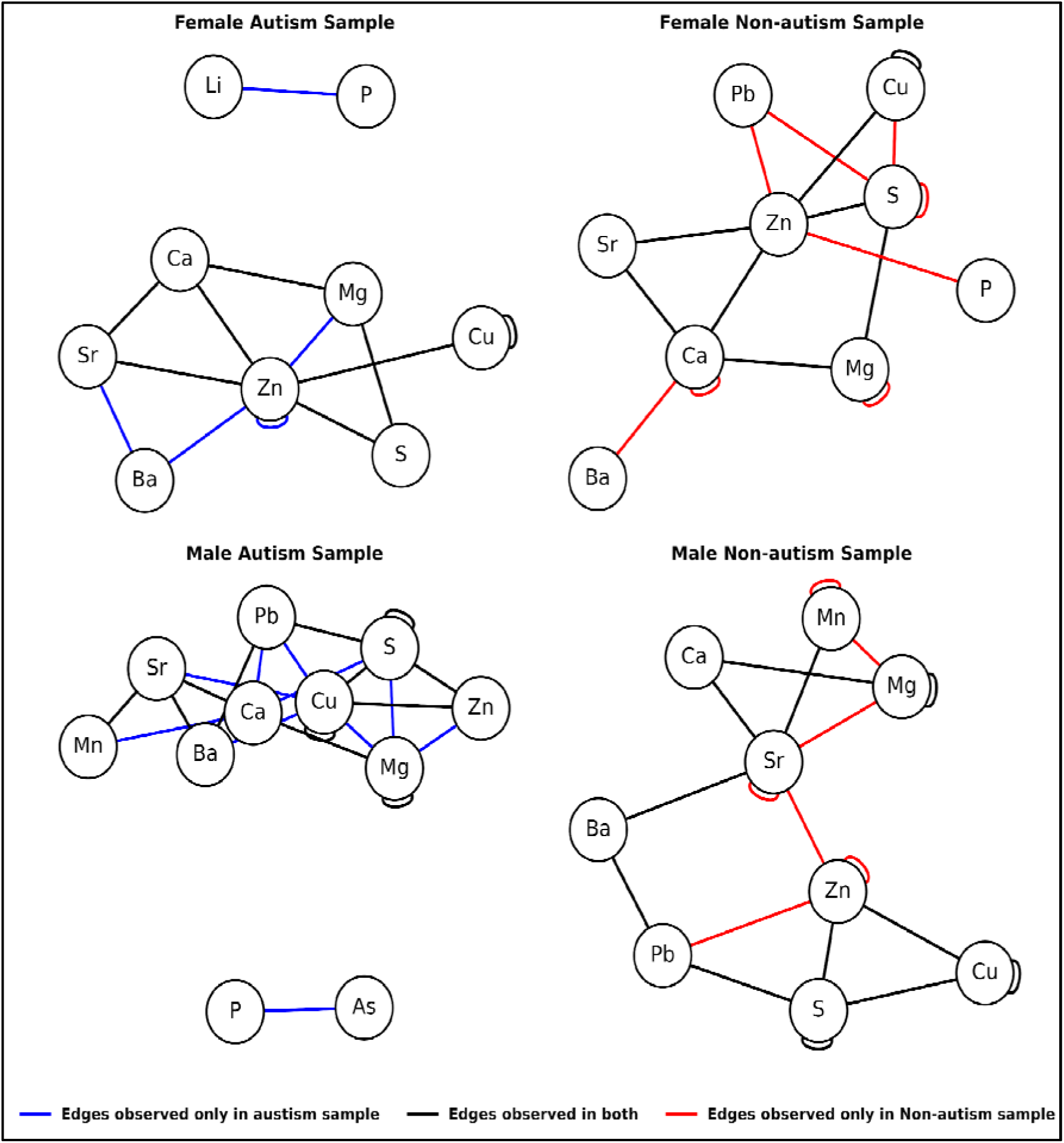
Illustration of interaction networks created using causal time series analysis between a sample with and without autism in our validation dataset, showing differences in connectivity among elements. Note that an edge between elements indicates a significant association (corrected for false discoveries at a significance level of 0.05) regardless of direction or strength. An edge in black indicates edges that exist in both the autism and non-autism samples; blue indicates edges that exist only in the autism sample; and red indicates edges that exist only in the non-autism sample.

The detailed modeling strategy is presented in Figure 4 (details are provided in Methods). During the training and stacking phase (see Figure 4), we found that the predictive signal was dispersed across multiple feature families and elements, indicating that the autism-associated signal is distributed across multiple biodynamic features rather than confined to a single representation. This is consistent with autism being a complex spectrum disorder that impacts multiple physiological systems rather than a condition arising from a single etiology. Our modelling strategy mimics such a distributed signal classification approach. In detail, base learners that met the training-phase criterion (*AUC* > 0.5) were advanced to the stacking stage, yielding 355 models for males and 305 for females from an initial 448. SHAP (SHapley Additive exPlanations) analyses of both the Stage 1 and Stage 2 meta-learners on the stacking set indicated distributed contributions rather than reliance on any single base model.^31^ In females, the highest-weighted contributors involved Li-Zn and Li-Ba biodynamics (using elemental network centrality features). In males, contributions were particularly diffuse, with prominent pairs such as P-Ba (using multiscale cross-entropy features), Mg-Cu, and, particularly for the Stage 1 meta-learner, Mg-Zn (using Catch22 features).^30,32^ This suggests that the discriminative signal for autism is distributed across multiple dynamical representations and multiple inter-element relationships.

**Figure 4:**
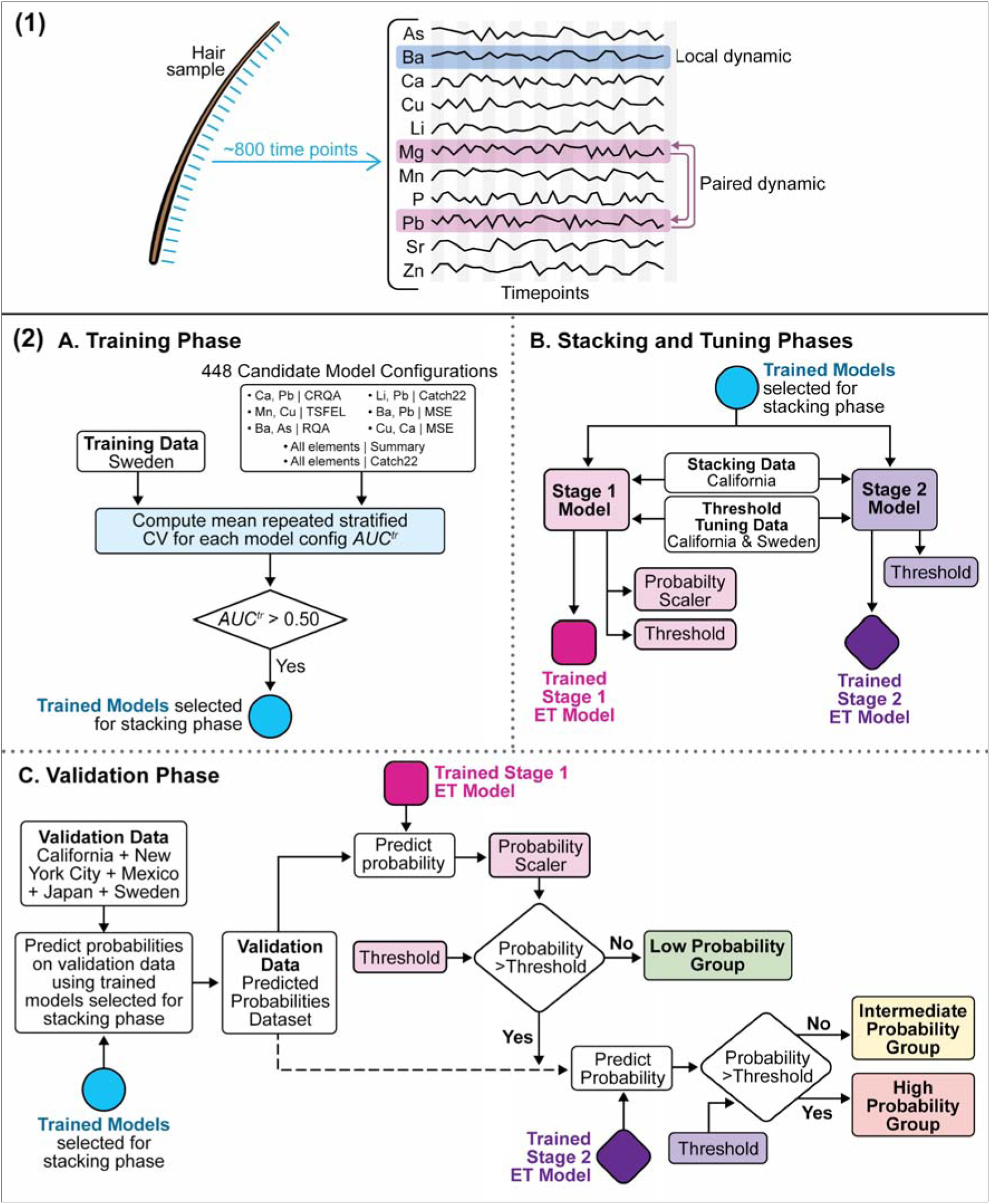
The figure presents an overview of (1) the temporality of elemental intensities from a hair strand, and (2) our modeling approach. Our machine-learning models explored both local and pair-wise dynamics in elemental intensities. (2A) denotes the training phase where models were trained using data from the RATSS cohort (Sweden), (2B) denotes the stacking phases where chosen models were stacked based on the CHARGE & MARBLES cohorts (California, USA), (2C-2D) denotes the validation phase where the final model is tested on multiple different cohorts, and once the models are finalized and tested on the multiple cohorts, threshold tuning is performed by optimizing sensitivity and specificity for classification into low-likelihood and high-likelihood groups respectively. ET: Extra Trees Classifier. *AUC^tr^*: training AUC

## Performance of the two-staged triage system

The sex-specific models were evaluated on independent validation cohorts comprising 580 samples from CHARGE/MARBLES (California), RATSS (Sweden), Mexico, JECS (Japan), and Seaver/PRISM (New York City). The age of the hair collection across these samples ranged from approximately 1 month to ∼37 years, with 91.2% of the samples collected below age 21. Both PRISM and JECS contributed only non-autism samples, as autism cases were either unavailable or excluded because DSM-5 criteria were not used during assessment (see Methods). This evaluation provided an assessment of generalizability across geographically and demographically diverse populations. Given the sequential triage system, to minimize false negatives and to ensure that those classified as low-likelihood were more likely to be controls, Stage 1 prioritized sensitivity and negative predictive value (NPV). Stage 2 focuses on specificity and positive predictive value (PPV), aiming to minimize false positives within the high-likelihood category. The results for the overall two-stage triage approach and each stage are summarized in Tables 1 and 2, respectively. Since autism is often characterized as a spectrum with diverse symptoms, our modeling strategy mimics it by creating more than two groups of autism likelihood. Instead of fully collapsing these three groups into merely two likelihood groups for reporting sensitivity and specificity metrics, we report the metrics hierarchically, which may better reflect the spectrum nature of the disorder. However, the full results (collapsing the three groups into two) are available in the supplemental appendix (Table S2), including sensitivity and specificity for each stage.

### Overall Two-Stage Model Performance

Figure 1B-C and Table 1 summarize the overall performance of the two-stage modeling approach, providing metrics across the three probability groups. When aggregating results across our two-stage approach, from the initial 580 validation set, our model stratified 113 (19.48%) samples as low-likelihood, 405 (69.83%) as intermediate-likelihood, and 62 (10.69%) as high-likelihood. Within each predicted group, the observed prevalence of autism also increased monotonically (low-likelihood: 4.42%, intermediate-likelihood: 21.98%, high-likelihood: 40.32%) from low- to high-likelihood groups. Assignment to progressively higher likelihood groups (low → intermediate → high) was associated with higher odds of autism, adjusted for sex (OR[95%CI]: 2.41[1.56-3.72]). Group-specific likelihood ratios also support our triage framework’s probability-based stratification performance, with higher likelihood ratios of 2.62 (95% CI:1.63-4.02) and 1.09 (95% CI:0.98-1.02) in the high- and intermediate-likelihood groups, respectively, compared to 0.18 (95% CI:0.15-0.22) in the low-likelihood group. Under sex-stratification, autism prevalence and group-specific likelihood ratios increased monotonically across likelihood groups (see Table 1). Odds ratios (OR[95%CI]) for males and females were 2.28[1.43-3.62] and 3.55[1.02-12.34] respectively.

### Low-Likelihood Stratification

In the first stage of low-likelihood stratification, the overall AUC of the model was 0.75 (95% CI: 0.70-0.79). For males, the model achieved an AUC of 0.64 (95% CI: 0.57-0.70, permutation-based p-value < 0.0001), with a sensitivity of 96% (95% CI: 0.91-0.99), an NPV (at 14% prevalence) of 96% (95% CI: 0.86-0.99), and a DLR- of 0.26. Because NPV is sensitive to population prevalence, at the general population autism rate of 3%, the NPV would be 99%. In females, the low-likelihood classifier showed comparable performance with an AUC of 0.69 (95% CI: 0.56-0.80, permutation-based p-value < 0.05) and a sensitivity of 92% (95% CI: 0.65-0.99). NPV at 14% and 3% prevalence were 96% (95% CI: 0.89-0.99) and 99% (95% CI: 0.94-1.00) respectively, with a DLR- of 0.24. The overall performance indicates that the model performed well as a low-likelihood classifier, minimizing false negatives while maintaining a high probability that those classified as “low-likelihood” are truly unaffected. Note that the overall AUC is influenced by the high predicted probability of male autism cases and the low predicted probability of female non-autism samples. We estimated the overall AUC directly from pooled predicted probabilities (rather than weighted averages) to facilitate thresholding and because most cohorts are not sufficiently well-balanced between males and females to justify sex-specific weighting. However, since the metrics (sensitivity and specificity) are directly obtained from separate sex-specific models, the overall AUC does not affect the results.

### Intermediate- and High-Likelihood Stratification

Next, in the second stage, we further classified the samples predicted by the stage 1 model to be not low-likelihood into intermediate- and high-likelihood groups. In males, we achieved an overall AUC of 0.56 (95% CI: 0.49-0.62, permutation-based p-value of 0.05), with a specificity of 86% (95% CI: 0.81-0.90), a PPV (at 14% prevalence) of 21% (95% CI: 0.14-0.30), and a DLR+ of 1.62 (see Table 2). In females, the second-stage model had an AUC of 0.61 (95% CI: 0.46-0.76, permutation-based p-value of 0.1), with a specificity of 96% (95% CI: 0.92-0.99) and a DLR+ of 2.08. PPV at 14% was 25% (95% CI: 0-0.65) and 6% (95%CI:0-0.26) at 3% autism prevalence.

### Sensitivity Analyses

Supplemental Tables S3-S6 summarize the stage-wise and overall performance of our likelihood stratification framework, testing the consistency of our results under validation cohort restrictions and aggregated bootstrapped stacking. (1) Validation Cohort Restrictions: Across all validation cohort restrictions, the association between predicted probability and autism status remained significant. Excluding the non-autism-only cohorts (JECS, PRISM) yielded a stronger association (OR[95% CI]: 3.1[1.86-5.17]) than in the original validation dataset (2.41[1.56-3.72]). Excluding samples 21 years and older yielded a similar association (2.34[1.51-3.64]) to the original validation dataset. Autism is frequently misclassified or misdiagnosed as intellectual disability due to overlapping symptoms like language delays and social communication deficits.^33,34^ Therefore, we conducted an additional sensitivity analysis on CHARGE/MARBLES samples with information on the Mullen Scales of Early Learning, which evaluate early intellectual development and motor skills. The odds were slightly elevated (2.63 [1.62-4.28]) when CHARGE/MARBLES samples with Mullen Scales of Early Learning scores below 70 were excluded. Stage-wise sensitivity and specificity also remained consistent under all cohort restriction analyses. (2) Bootstrapped Aggregated Stacking: Bootstrap aggregation of the stacking dataset yielded an AUC of 0.72 (95% CI: 0.67-0.77) for low-likelihood stratification and 0.70 (95% CI: 0.65-0.75) for high/intermediate-likelihood stratification, indicating the stability of our ensemble models. (3) Threshold Variations: We also established the robustness of the threshold tuning by demonstrating how performance varies around the neighborhood of the estimated thresholds (see supplemental appendix, Figure S1). (4) Location - Based Discrimination: Location-based discriminative power in stage 1 was broadly consistent across geographic cohorts with no evidence for a cohort effect. Within the US, AUCs ranged from 0.74 (95% CI: 0.62-0.83) for New York to 0.77 (95% CI: 0.67-0.84) for California. Non-US cohorts showed similar stability, with Mexico at 0.78 (95% CI: 0.57-0.91) and RATSS (Sweden) at 0.71 (95% CI: 0.60-0.80) within the same range.

## DISCUSSION

Here, we introduce a novel biochemical hair-based machine-learning tool designed to stratify autism probability in children as young as one month. In our proof-of-concept results from multiple populations, the likelihood stratification tool achieved a high sensitivity of 96% in stratifying non-autism controls as low probability, whereas the second-stage model achieved a specificity of 90% in identifying a high-probability group among participants not triaged as low probability. The likelihood ratios for autism diagnosis reinforce this interpretation, with a monotonic increase observed in the low-, intermediate-, and high-probability groups (likelihood ratios = 0.18, 1.09, and 2.62, respectively). These metrics suggest that this likelihood stratification tool could enable healthcare providers to efficiently triage individuals on long autism assessment waiting lists. Our approach moves the current health care systems from a first-come-first-served model to one where access to clinical services is prioritized based on objectively measured autism probability, thereby improving timely access to clinical services for those most in need, preserving clinician time for those more likely to be on the autism spectrum, and improving the cost-effectiveness of autism service networks. Because this biochemical stratification tool can be implemented in the first year of life, it can also reduce the age at initiation of therapy, which has been shown to have greater benefits when started earlier.^35^ Furthermore, the non-invasive nature of hair sampling enhances the assay’s clinical utility, facilitating broader accessibility and compliance.

Multiple potential biomarkers of autism are currently being extensively studied, ranging from genetic (autism polygenic score), epigenetic (histone modification changes), immunologic (brain autoantibodies, folate receptor alpha autoantibody, and cytokines), metabolic (altered purine metabolism, ATP-related purinergic signalling, fatty acid oxidation, as well as change in metabolic pathways related to energy, neurotransmitters, amino acids, and mitochondrial dysfunction), neurological (Morphology and diffusion tensor imaging, MRI, visual attention), and autonomic (heart rate variability, dentine-based marker).^36–50^ Although biomarker research on autism is active, most promising biomarkers are still in their early stages of development. Furthermore, most are not yet suitable for large-scale clinical implementation due to either their potential invasive nature or the difficulty of implementation. Almost all studies on the effectiveness of biomarkers are based on either correlational or association-type studies; therefore, although such results can yield robust statistical inferences, generalizability and the identification of predictive patterns remain challenging.^51^ Finally, the plausibility of varied autism subtypes renders such association-based results difficult to generalize to external, unseen samples.

Bae et al. recently introduced a two-stage autism stratification tool using parent-child interaction audio and screening data in children aged 18-48 months.^52^ In contrast, our framework examines biochemical signals and extends autism likelihood stratification to early infancy, including infants as young as 1 month. Similar to Bae et al., our framework is not based on a single modelling layer but instead applies a sequential architecture that refines classification in a stepwise manner, somewhat analogous to a shallow tree-based model such as a gradient-boosted decision tree. A model designed like this not only leverages predictive feature ensembling but also harnesses the synergistic power of ensembling two relatively weaker predictive models trained to correct the “residuals” of the previous model.

Moreover, a sequential structure such as this is well-suited to the incorporation of diverse data modalities. As measurement technologies continue to improve, one might, for example, in the second layer, incorporate additional molecular features that contribute information that is complementary and potentially independent to the elemental signals used here. Although including multiple layers essentially makes the machine learning tool more data-hungry, the modularity of this layered architecture ensures that future improvements can be made without completely altering the initially developed core layers.

This work builds on our previous proof-of-concept associative analyses, which identified a few features with a high degree of association using a much smaller sample.^53^ However, a strong association does not necessarily imply a firm prediction.^54^ In this paper, we implemented a full ML architecture using a much larger dataset to improve the potential for generalization to unseen data. We leveraged state-of-the-art techniques to model elemental biodynamic features, potentially capturing higher-order interactions among elements.^24^ This framework also included both internal cross-validation and external validation using independent datasets from disparate and geographically distinct cohorts. Central to our approach was the use of a layered, multi-tiered model-stacking strategy, in which candidate models had to meet a pre-specified statistical performance threshold to advance to the next phase of modelling. Moreover, to further mitigate the risk of model overfitting and spurious findings, we conducted detailed sensitivity analyses.

Overall, we observed slightly lower performance metrics in males compared to females. This discrepancy may reflect underlying differences in the phenotypic expression of autism between sexes. Some research suggests that autistic females, compared to their male counterparts, might require presenting with more autistic behaviors to be diagnosed.^55,56^ It is also likely that given some autism phenotypes in females differ from those in males, such a discrepancy leads to underdiagnosis and particularly later diagnoses associated with masking of challenges, internalization of symptoms, and biases in diagnostic practices. Younger females with autism are often more motivated to learn or mimic socially acceptable behaviors, masking their difficulties and making it harder to recognize their autism-related phenotype.^57^

There are several strengths of this work worth noting. Foremost, we have demonstrated that our likelihood stratification model can effectively stratify autism likelihood in children as young as 1 month of age, setting a new standard for autism screening. We collected hair samples from participants spanning a wide age range, enabling us to assess the stability of our model across developmental periods. We have tested our model across multiple geographic regions, under varying degrees of outcome misclassification, to mimic plausible real-world clinical settings and demonstrate its generalizability across different environmental exposures, diets, socioeconomic factors, ethnicities, and other variables. The relatively large sample size in our study enabled us to conduct multiple sensitivity analyses. This provides preliminary evidence that this likelihood-stratification model could potentially serve as an additional autism screening tool during children’s regular visits to their pediatrician. Collection of hair samples is easy and non-invasive, requiring only a small amount of hair, making this diagnostic test readily implementable across various settings.

This study and the proposed li stratification tool also have limitations that need to be addressed. First, and most importantly, the machine learning tool is based solely on data on elemental metabolism. In the future, it will be necessary to develop a stratification layer that includes additional biochemical markers. Second, some subgroup sizes, particularly among female cases, were modest, which limits the precision of subgroup-specific estimates. The imbalance in sex distribution reflects the historically higher diagnosis rates of autism in males.^58^ Future work involving larger datasets and newer techniques may offer greater sensitivity to weak signal patterns and warrant further exploration. Moreover, this would provide additional information that may also help reduce the size of the intermediate-likelihood group. Third, while the study spans multiple countries and diverse populations, capturing variation in ancestry, socioeconomic background, environmental exposures, and healthcare access, additional sites would enhance the generalizability of our findings.^59^ Additional efforts will be required to identify biomarkers of phenotypes and comorbidities that enable individualized therapies to be delivered at an earlier age. Fourth, the Stage 2 model was developed after the Stage 1 model had been developed and evaluated on the validation set. Although this sequential approach reflects the intended use of the framework, future work could explore simultaneous pre-specification and development of both stages within a unified training strategy. Lastly, a future goal of our work may include stratification layers for gene-environmental interactions to capture additional synergistic effects that might be missed with a single data source.

Overall, we show that analyzing elemental biodynamics using single hair strands may provide an objective approach to stratifying autism likelihood earlier than is possible with current clinical observational tools. Given the long waiting times and highly resource-intensive diagnostic odyssey families currently experience, this likelihood-stratification could help clinicians triage patients based on an objective measure of autism risk, ensuring that high-likelihood cases are not missed.

## MATERIALS AND METHODS

The combined sample (N = 1697) used in this analysis was collected from diverse study populations across multiple countries, including the United States, Sweden, Mexico, and Japan. Details on the IRB can be found in the Ethical Approval Statement. A schematic of the study samples and corresponding cohorts, along with their use in modelling, is shown in Figure 2.

### Study Population

This study included a total of 1697 samples across six cohorts: CHARGE (n = 825) and MARBLES (n = 287) from California, RATSS from Sweden (n = 306), Seaver (n = 39) and PRISM (n = 84) cohorts from New York, JECS from Japan (n = 110), and a cohort from Mexico (n = 46). Only one hair strand was used for each participant (see supplementary appendix S2 for more information). Table 3 summarizes the demographic and elemental characteristics within each cohort. In CHARGE, males comprised 77.1% of all samples, with a median age at the time of hair collection of 36 months. In MARBLES, participants were younger (median 12.5 months) at the time of hair collection, with a more balanced sex distribution overall (55.8% males), although cases were disproportionately male (74.2%). Among the non-autism group, 51% and 20% had developmental delays, in CHARGE and MARBLES, respectively. RATSS participants were older at the time of hair collection (median 168 months), with 55.6% males. The Seaver cohort had a median age of 72 months and also showed a predominance of males among cases. The Mexico cohort had a median age of 35.5 months at the time of hair collection and was predominantly male (73.9%). The PRISM cohort consisted of younger participants (compared to the other NYC cohort), with a median age of 48 months at the time of hair collection and 61% male. Children from RPISM were non-autism participants from the general population (no children were referred for a specialist diagnostic assessment). Overall, the youngest participants were from JECS (all non-autism controls), with a median age of hair collection of 1 month and a balanced sex distribution (50% males). Average ablated hair lengths were highly consistent across CHARGE, MARBLES, and Seaver (∼1.19 cm), somewhat lower in PRISM and Mexico (∼1.16 cm) and JECS (∼1.14 cm), and substantially shorter in RATSS (∼0.91 cm). Full graphical comparisons of measured elements across subgroups are provided in the supplementary appendix, Figures S2-S12.

### CHARGE and MARBLES (California, USA)

Data were collected from the Childhood Autism Risks from Genetics and Environment (CHARGE) and the Markers of Autism Risk in Babies—Learning Early Signs (MARBLES) studies, recruited in California at the MIND Institute, University of California, Davis.^60,61^ Informed consent was obtained before participation and data collection in each study. The protocols for the CHARGE and MARBLES cohorts have been previously described.^60,61^ CHARGE began recruitment in 2003. It is a case-control study that includes children receiving services for autism or developmental delay from Regional Centers that contract with the California Department of Developmental Services and children recruited from the general population from birth records, frequency-matched to the autism cases on sex, geographic region, and age. Those entering with a prior autism diagnosis confirmed by both the Autism Diagnostic Observation Schedule Second Edition (ADOS-2) and Autism Diagnostic Interview—Revised (ADI-R). All other participants were screened with the Social Communication Questionnaire (SCQ).^62^ Those children whose SCQ score was above 14 were further administered the ADI-R and ADOS-2.^60^ CHARGE samples used in this study include either hair saved by parents from the child’s first haircut or hair collected at the child’s clinic visit; thus, some children were diagnosed after, and some before, the hair sample collection. For CHARGE, the non-autism group included all children who did not meet the diagnostic criteria for autism, encompassing those classified as typically developing, developmentally delayed, or those referred to the study with either autism or developmental delay who did not meet criteria for autism diagnosis or developmental delay classification. MARBLES is an elevated-likelihood birth cohort recruiting pregnant women who either have a child or a first-degree relative with autism. Recruitment began in 2006. Children were evaluated at 3 years of age using the ADOS-2. For MARBLES samples, hair was typically collected between 3 and 24 months during a clinic visit, and an autism diagnosis was made around age 36 months. For MARBLES, the non-autism group included all children who did not meet the diagnostic criteria for autism, including those classified as typically developing and those classified as not typically developing (e.g., developmentally delayed). Clinicians administering the CHARGE and MARBLES instruments are trained and have formally demonstrated research reliability on the ADOS. Both CHARGE and MARBLES also assessed children’s cognitive development using the Mullen Scales of Early Learning (MSEL).^63^ See supplementary appendix Section S2 for additional details.

### Seaver and PRISM (New York, USA)

Data was collected from the Seaver Autism Center at the Icahn School of Medicine at Mount Sinai, New York City.^64^ This center receives referrals related to autism from the surrounding area. Beginning in 2016, families already engaged with Seaver Autism Center were approached if they had a child with a confirmed autism diagnosis and at least one sibling without autism, with hair samples typically collected from both siblings. Diagnoses were supported by the ADOS-2 and ADI-R, along with a clinical evaluation conducted by a board-certified child and adolescent psychiatrist or a licensed clinical psychologist, in accordance with the DSM-5 diagnostic criteria. We further included general population-based participants drawn from the PRogramming of Intergenerational Stress Mechanisms (PRISM) study, which recruited children whose mothers received prenatal care at Mount Sinai, into a longitudinal study designed to investigate the influence of psychological stress and other environmental influences on child development.^65^ The child version of the Autism Spectrum Quotient (AQ-10) was completed for all children, and no children were referred for a specialist diagnostic assessment. In this paper, these children were treated as non-autism participants.^66^

### RATSS (Sweden)

Data was also collected from the Roots of Autism and ADHD Twin Study Sweden (RATSS).^67^ Participants (twins) were recruited for this study between 2011 and 2016. Participants included both concordant and discordant twin pairs: those in which one or both twins met the criteria for autism, as well as pairs in which neither twin did. Details on participant recruitment have been previously published.^68^ Autism screening was conducted at age nine. DSM-5 criteria were used for autism diagnosis, and results were corroborated using the ADI-R and ADOS-2. RATSS included detailed data on ADHD and intellectual disability diagnoses, as well as psychiatric comorbidities. Given the availability of this data, participants with any intellectual disability or other psychiatric comorbidities were excluded, since children with autism have a higher likelihood of being misdiagnosed with intellectual disability.^33,69^ For RATSS, the non-autism group comprised participants who did not meet the diagnostic criteria for autism, whether typically developing or with other neurodevelopmental conditions.

### JECS (Japan)

Data were additionally obtained from the Japan Environment and Children’s Study (JECS), a large-scale, nationwide birth cohort designed to investigate environmental influences on children’s health.^70^ Recruitment was conducted across 15 regional centers throughout Japan. Hair samples were collected from infants at approximately one month of age and archived at the JECS biorepository under standardized conditions. Autism diagnosis was carried out around age four. For the present study, 220 children were randomly selected from among participants who had completed the three-year follow-up questionnaire. Of these, 110 participants were treated as non-autism samples (using AQ-10) and included in the validation set (to determine whether including a geographically diverse set of non-autism samples affected the accuracy of our models). Autistic individuals identified within JECS were excluded due to concerns about diagnostic reliability, particularly the lack of DSM-5 consensus with ADI-R and ADOS-2. While this precluded balanced autism/non-autism analyses within JECS, inclusion of non-autism samples broadened the demographic and geographic scope of the validation dataset.

### Mexico City (Mexico)

Recruitment was conducted through a campaign by Domus Institute of Autism and Iberoamericana University, focusing on families with undiagnosed children younger than 7 years, and with the following inclusion criteria: children with developmental delays or signs of autism, or children with a sibling or immediate family member diagnosed with autism. Children accompanied by their parents provided signed informed consent and hair specimens at the study visit (Domus Institute of Autism). Autism diagnostic status was determined based on clinical evaluations conducted by trained, standardized psychologists within the same week as hair collection, from August 2024 to May 2025. During the evaluation, clinicians systematically coded and scored observed behaviors according to standardized ADOS-2 protocols. Importantly, the ADOS-2 was not used as a stand-alone diagnostic tool; its results were integrated with developmental history, family interviews, clinical observation, and complementary assessments (including ADI-R and cognitive and adaptive evaluations) to reach a reliable and comprehensive clinical diagnosis of autism. For this cohort, the non-autism group consisted of participants who did not meet the diagnostic criteria for autism.

## LABORATORY PROTOCOLS AND SIGNAL PROCESSING

Elemental measurements were acquired using laser ablation-inductively coupled plasma-mass spectrometry (LA-ICP-MS) under standardized daily tuning and ongoing quality control procedures across participating laboratories. Calibration and performance monitoring were conducted using certified reference materials and laboratory-specific quality control materials, and analytical runs were released only after meeting established QA/QC criteria. The majority of samples were analyzed at *LinusBio* clinical laboratory (CHARGE, MARBLES, Mexico, and a subset of RATSS), with a minority analyzed at Icahn School of Medicine at Mount Sinai (Seaver, PRISM, JECS, and a subset of RATSS), employing comparable sample preparation, instrument analysis, and normalization procedures. Details on laboratory sample preparation, instrumentation, and analysis, and QA/QC procedures are provided in Supplementary Section S1. Raw ion counts for each element per ablation time point are used to construct the elemental time series. To correct for hair-density/ablation variability and instrumental drift, we applied pointwise sulfur (S) normalization. At each time point, we computed the ratio of the element count to the Sulfur count (e.g., Cu/S). Sulfur itself was not normalized. Downstream feature extraction and modelling was carried out using 12 elements: magnesium (Mg), phosphorus (P), calcium (Ca), copper (Cu), zinc (Zn), strontium (Sr), barium (Ba), lead (Pb), lithium (Li), manganese (Mn), Arsenic (As) and Sulphur (S).

## STATISTICAL METHODS

On a sample of individuals drawn from the RATSS, CHARGE, and MARBLES study cohorts, we trained (and stacked) machine learning models using temporal biodynamic features computed from multivariate elemental time series. We assumed that potential heterogeneity in autism etiologies may manifest as differences in elemental biodynamics.^71,72^ To capture diverse aspects of elemental biodynamics, we used eight feature extraction techniques, encompassing both univariate features (e.g., entropy-complexity) computed for individual elements and bivariate features (e.g., cross recurrence quantification analysis [CRQA] and elemental network centrality) computed across element pairs.^73–75^ Full details about these techniques are available in Supplementary Section S3. To account for potential sexual dimorphic heterogeneity in autism diagnosis, we stratified the models by sex. Although the mechanisms underlying sex differences in the etiology of autism are not fully understood, prior research suggests differential phenotypic signatures.^57,76,77^ This stratified approach enabled modelling within more clinically homogeneous subgroups.

### Modelling Framework

We developed a two-stage modelling framework designed to sequentially stratify subjects into three likelihood groups: low, intermediate, and high. The first stage identifies samples at low likelihood of autism, with the second stage further classifying the remaining samples into intermediate and high-likelihood groups. All modelling procedures were constructed separately for each sex. The framework (see Figure 4) consisted of (1) a Training Phase, in which candidate models were trained and screened based on cross-validated performance. (2) a Stacking Phase, in which these filtered candidate models were combined to create two different model ensembles for low and high-likelihood classification using stacked generalization.^78^ A final external validation was conducted on held-out datasets, where the final performance was reported. Here we provide a brief overview. See supplementary appendix for more description (Section S3-S4).

#### Training Phase

We constructed a candidate model space by exhaustively combining two components: (1) predefined elemental subsets, including all 55 unique pairs among 11 elements excluding Sulphur, and a full model using all 12 elements; and (2) 8 time series feature extraction techniques. This resulted in 448 candidate model combinations per sex. For each candidate combination (e.g., [Ca, Pb | CRQA]), we fitted an elastic-net-penalized logistic regression. Performance was evaluated using repeated stratified cross-validation. The mean cross-validated area under the curve (AUC) was computed for each candidate model combination (*AUC^tr^*), and only those with *AUC^tr^*>0.5 were advanced to the stacking phase.

#### Stacking Phase

Candidate models that passed training-phase criteria were combined via stacked generalization.^78^ An independent stacking dataset was used to fit two separate meta-learners that ingest the base learners’ predicted probabilities as features. For each meta-learner, we used an *ExtraTrees* classifier (also called Extremely Randomized Trees).^79^ To assess the relative contribution of the individual base models within the stacked ensemble, we applied SHAP analyses (tree-SHAP) to the stacking dataset, attributing the meta-learner’s predictions to each base-model feature.^80^ The stacking process was carried out within each sex.

#### Stage 1 Model (Low-likelihood Classification)

The Stage 1 model was designed to identify a low-likelihood group while maintaining high sensitivity (minimised false negative rate). After training the stage 1 meta-learner, predicted probabilities were generated on a dedicated tuning set (see Figures 2 and 3). The probability threshold was selected to meet performance targets for high sensitivity and negative predictive value (NPV) (both >90%). Among the range of probability values that meet these target criteria, the 10th percentile probability threshold was selected, and robustness was evaluated using sensitivity analysis. Individuals with predicted probabilities below this threshold were assigned to the low-likelihood category. Those not assigned to the low-likelihood group proceeded to Stage 2.

#### Stage 2 Model (High-likelihood Classification)

A second meta-learner model was fitted on the stacking data and then applied to individuals not classified as low likelihood to further stratify them into high- and intermediate-likelihood groups. The Stage 2 model used the same modeling architecture as Stage 1, including elastic-net-penalized logistic regression base learners and an ExtraTrees meta-learner with similar class weighting and number of trees. However, to introduce a slight differential learning mechanism, improve stability, and reduce overfitting, the Stage 2 meta-learner was regularized by limiting tree depth (*max_depth*=5), whereas the Stage 1 model allowed fully expanded trees (*max_depth*=None). These meta-learners were trained independently for Stage 1 and Stage 2, but shared the same underlying feature space (the probabilistic outputs of the base learners). To define a high-likelihood threshold, out-of-fold predicted probabilities were generated from the stacking dataset using stratified cross-validation. The probability threshold was prespecified as the 90th percentile of out-of-fold predicted probabilities among controls within the stacking dataset, corresponding to a target specificity of approximately 90%. This threshold was prespecified before external validation. Individuals exceeding this threshold were classified as high likelihood, while remaining individuals were classified as intermediate likelihood.

#### Final Testing and Evaluation Metrics

The final models (Stage 1 and 2) for each sex were validated using additional samples from the RATSS, CHARGE, and MARBLES cohorts as well as an aggregate of four independent external cohorts that had not been used for model training, stacking, or tuning. These included: SEAVER and PRISM (New York, USA), samples from Mexico City, and the JECS cohort from Japan (see Figure 2). The inclusion of these geographically and demographically distinct populations enabled evaluation of generalizability.

For Stage 1, model performance was primarily assessed using AUC, sensitivity, and NPV. Sensitivity in this context was defined as the proportion of cases not assigned to the low-likelihood category. For Stage 2, model performance among participants not classified as low likelihood was assessed using AUC, specificity, and positive predictive value (PPV). Specificity in this context was defined as the proportion of controls not assigned to the high-likelihood category. Given the sequential triage system, Stage 1 prioritised sensitivity and NPV to minimise false negatives and to ensure that those classified as low likelihood had minimal probability of autism. Stage 2 emphasised specificity and PPV, with the goal of minimising false positives and enriching autism cases within the high-probability category. Additionally, we report the negative and positive diagnostic likelihood ratios (DLR-, DLR+) for stages 1 and 2, respectively. For both stages, predictive values (NPV and PPV) were estimated under prevalences of 3% (general population) and 14% (elevated-likelihood population)^81,82^. For AUC estimates, permutation-based p-values were estimated using 5000 random-label permutations, and 95% confidence intervals were estimated using bootstrap resampling with 1000 iterations. For the overall triage framework, we reported the distribution of participants and the prevalence of autism across low-, intermediate-, and high-probability groups, as well as group-specific likelihood ratios (LR = P(group|autism)/P(group|non-autism)) and odds ratios for autism across ordered likelihood groups.

#### Training, Stacking, Tuning, and Validation Subsets

For model training, 138 RATSS samples (42 cases) were used. A further 571 samples (339 from CHARGE and 232 from MARBLES) were allocated for stacking. An additional 546 samples were used as a tuning set (for the Stage 1 model), comprising 408 non-autism samples from CHARGE and MARBLES, and the 138 RATSS samples used in the model training phase. This construction, by design, emphasized the classification of samples into low-likelihood. To introduce variation from the stage 1 model, the stacking set was reused as a tuning set for stage 2. Finally, 580 samples were reserved for independent validation. These samples were collected from multiple cohorts to provide geographic and demographic diversity, including CHARGE/MARBLES (California), RATSS (Sweden), Mexico, JECS (Japan), and New York (Seaver and PRISM). Figure 2 shows the schematic of cohorts at various modeling phases and validation. These subgroups varied in size and prevalence of autism (Table S1, supplementary appendix).

#### Sensitivity Analyses

To evaluate the robustness of our modelling framework, we performed sensitivity analysis of our sequential models: (1) by introducing restrictions in the validation sample; (2) with an aggregated bootstrapped stacking stage; (3) under threshold variations; and (4) testing location-based discriminative ability at the stage 1 level. First, restrictions were introduced in the validation samples by excluding: (i) both non-autism only cohorts (JECS [Japan] and PRISM [New York]); (ii) children from California validation data with MSEL (Mullen Scales of Early Learning) score below 70;^83,84^ and (iii) participants with age of hair collection at and above 21 years. Second, for stacking, we tested the stability of the stage 1 and 2 meta-learners by repeatedly resampling the stacking dataset using a bootstrap aggregation approach (100 bootstrap samples).^85^ Third, for thresholds, we assessed the robustness of the threshold-tuning procedure by examining how sensitivity/specificity and NPV/PPV vary across all probability thresholds that met our predefined performance criteria for both the stage 1 and stage 2 models (see supplementary appendix, Figure S1). Lastly, we examine our models’ discriminative ability for each validation location separately, using their stage 1 AUCs.

## Ethical Approval Statement

This current study is approved by the Mount Sinai IRB. The CHARGE and MARBLES study protocols were approved by the University of California at Davis and the State of California Institutional Review Boards. For SEAVER and PRISM, the present study was approved by the Mount Sinai IRB, and written informed consent was obtained from all participants, their parents, or guardians. The RATSS study was approved by the Swedish Regional Ethical Review Board. The protocol for JECS was reviewed and approved by the IRB of the Japanese Ministry of the Environment’s on Epidemiological Studies and by the Ethics Committees of all participating institutions. All participating women provided written informed consent. The study in Mexico City was approved by the ethical review board of IberoAmericana University. For all study populations included in this manuscript, informed consent was obtained after the nature and possible consequences of the studies were explained.

## Data Availability

Clinical data from this study cannot be posted to a publicly accessible forum, as they contain private health information. However, they will be available from authors at reasonable request, subject to appropriate clearances from research governance authorities at each participating study institution.

## References

1. Zeidan J, Fombonne E, Scorah J, et al. Global prevalence of autism: A systematic review update. Autism research 2022; 15(5): 778–90.

2. Shaw KA. Prevalence and early identification of autism spectrum disorder among children aged 4 and 8 years—Autism and Developmental Disabilities Monitoring Network, 16 Sites, United States, 2022. MMWR Surveillance Summaries 2025; 74.

3. Nasir AK, Strong-Bak W, Bernard M. Diagnostic evaluation of autism spectrum disorder in pediatric primary care. Journal of primary care & community health 2024; 15: 21501319241247997.

4. Gordon-Lipkin E, Foster J, Peacock G. Whittling down the wait time: exploring models to minimize the delay from initial concern to diagnosis and treatment of autism spectrum disorder. Pediatric Clinics of North America 2016; 63(5): 851.

5. Bölte S, Girdler S, Marschik PB. The contribution of environmental exposure to the etiology of autism spectrum disorder. Cellular and molecular life sciences 2019; 76(7): 1275–97.

6. Havdahl A, Niarchou M, Starnawska A, Uddin M, Van Der Merwe C, Warrier V. Genetic contributions to autism spectrum disorder. Psychological medicine 2021; 51(13): 2260–73.

7. Lyall K, Croen L, Daniels J, et al. The changing epidemiology of autism spectrum disorders. Annual review of public health 2017; 38: 81–102.

8. Modabbernia A, Arora M, Reichenberg A. Environmental exposure to metals, neurodevelopment, and psychosis. Current opinion in pediatrics 2016; 28(2): 243–9.

9. Midya V, Alcala CS, Rechtman E, et al. Machine learning assisted discovery of interactions between pesticides, phthalates, phenols, and trace elements in child neurodevelopment. Environmental Science & Technology 2023; 57(46): 18139–50.

10. Seabra CM, Aneichyk T, Erdin S, et al. Transcriptional consequences of MBD5 disruption in mouse brain and CRISPR-derived neurons. Molecular autism 2020; 11(1): 45.

11. Curtin P, Austin C, Curtin A, et al. Dysregulated biodynamics in metabolic attractor systems precede the emergence of amyotrophic lateral sclerosis. PLoS Computational Biology 2020; 16(4): e1007773.

12. Arora M, Giuliani A, Curtin P. Biodynamic interfaces are essential for human–environment interactions. Bioessays 2020; 42(11): 2000017.

13. Huo M, Zhang J, Huang W, Wang Y. Interplay among metabolism, epigenetic modifications, and gene expression in cancer. Frontiers in Cell and Developmental Biology 2021; 9: 793428.

14. Malavolta M, Piacenza F, Basso A, Giacconi R, Costarelli L, Mocchegiani E. Serum copper to zinc ratio: Relationship with aging and health status. Mechanisms of ageing and development 2015; 151: 93–100.

15. Djoko KY, Cheryl-lynn YO, Walker MJ, McEwan AG. The role of copper and zinc toxicity in innate immune defense against bacterial pathogens. Journal of Biological Chemistry 2015; 290(31): 18954–61.

16. Billman GE. Homeostasis: the underappreciated and far too often ignored central organizing principle of physiology. Frontiers in physiology 2020; 11: 200.

17. Arora M, Austin C, Sarrafpour B, et al. Determining prenatal, early childhood and cumulative long-term lead exposure using micro-spatial deciduous dentine levels. PloS one 2014; 9(5): e97805.

18. Austin C, Smith TM, Bradman A, et al. Barium distributions in teeth reveal early-life dietary transitions in primates. Nature 2013; 498(7453): 216–9.

19. Cheng N, Rho JM, Masino SA. Metabolic dysfunction underlying autism spectrum disorder and potential treatment approaches. Frontiers in molecular neuroscience 2017; 10: 34.

20. Frye RE. Mitochondrial dysfunction in autism spectrum disorder: unique abnormalities and targeted treatments. Seminars in pediatric neurology; 2020: Elsevier; 2020. p. 100829.

21. Wu Q, Ren Q, Meng J, Gao W-J, Chang Y-Z. Brain iron homeostasis and mental disorders. Antioxidants 2023; 12(11): 1997.

22. Jeyasingh PD, Goos JM, Thompson SK, Godwin CM, Cotner JB. Ecological stoichiometry beyond redfield: an ionomic perspective on elemental homeostasis. Frontiers in Microbiology 2017; 8: 722.

23. Austin C, Curtin P, Curtin A, et al. Dynamical properties of elemental metabolism distinguish attention deficit hyperactivity disorder from autism spectrum disorder. Translational psychiatry 2019; 9(1): 238.

24. Midya V, Bello G, Andrew AS, Re DB, Stommel EW, Arora M. Dysregulation of hair-strand-based elemental biodynamics in amyotrophic lateral sclerosis. Ebiomedicine 2025; 119.

25. Braddock B, Turner K, Kumarason K, et al. Improving Access to Autism Evaluation: A Coordinated Developmental-Behavioral Pediatrics and Pediatric Neurology Diagnostic Pathway. Missouri Medicine 2024; 121(3): 225.

26. Brian JA, Zwaigenbaum L, Ip A. Standards of diagnostic assessment for autism spectrum disorder. Paediatrics & child health 2019; 24(7): 444–51.

27. Zbilut JP, Webber Jr CL. Recurrence quantification analysis. Wiley encyclopedia of biomedical engineering 2006.

28. Wallot S, Leonardi G. Analyzing multivariate dynamics using cross-recurrence quantification analysis (crqa), diagonal-cross-recurrence profiles (dcrp), and multidimensional recurrence quantification analysis (mdrqa)–a tutorial in r. Frontiers in psychology 2018; 9: 2232.

29. Flood MW, Grimm B. EntropyHub: An open-source toolkit for entropic time series analysis. PloS one 2021; 16(11): e0259448.

30. Lubba CH, Sethi SS, Knaute P, Schultz SR, Fulcher BD, Jones NS. catch22: CAnonical Time-series CHaracteristics: Selected through highly comparative time-series analysis. Data mining and knowledge discovery 2019; 33(6): 1821–52.

31. Chen H, Covert IC, Lundberg SM, Lee S-I. Algorithms to estimate Shapley value feature attributions. Nature Machine Intelligence 2023; 5(6): 590–601.

32. Jamin A, Humeau-Heurtier A. (Multiscale) cross-entropy methods: A review. Entropy 2019; 22(1): 45.

33. Thurm A, Farmer C, Salzman E, Lord C, Bishop S. State of the field: Differentiating intellectual disability from autism spectrum disorder. Frontiers in psychiatry 2019; 10: 526.

34. Etyemez S, Esler A, Kini A, et al. The role of intellectual disability with autism spectrum disorder and the documented cooccurring conditions: A population-based study. Autism Research 2022; 15(12): 2399–408.

35. Guthrie W, Wetherby AM, Woods J, et al. The earlier the better: An RCT of treatment timing effects for toddlers on the autism spectrum. Autism 2023; 27(8): 2295–309.

36. de Wit MM, Morgan MJ, Libedinsky I, et al. Systematic review and meta-analysis: Phenotypic correlates of the autism polygenic score. JAACAP open 2025.

37. Sun W, Poschmann J, Del Rosario RC-H, et al. Histone acetylome-wide association study of autism spectrum disorder. Cell 2016; 167(5): 1385–97. e11.

38. Jensen AR, Lane AL, Werner BA, McLees SE, Fletcher TS, Frye RE. Modern Biomarkers for Autism Spectrum Disorder: Future Directions: AR Jensen et al. Molecular diagnosis & therapy 2022; 26(5): 483–95.

39. Saghazadeh A, Ataeinia B, Keynejad K, Abdolalizadeh A, Hirbod-Mobarakeh A, Rezaei N. A meta-analysis of pro-inflammatory cytokines in autism spectrum disorders: Effects of age, gender, and latitude. Journal of psychiatric research 2019; 115: 90–102.

40. Frye RE, Slattery JC, Quadros EV. Folate metabolism abnormalities in autism: potential biomarkers. Biomarkers in medicine 2017; 11(8): 687–99.

41. Lingampelly SS, Naviaux JC, Heuer LS, et al. Metabolic network analysis of pre-ASD newborns and 5-year-old children with autism spectrum disorder. Communications Biology 2024; 7(1): 536.

42. Rose S, Niyazov DM, Rossignol DA, Goldenthal M, Kahler SG, Frye RE. Clinical and Molecular Characteristics of Mitochondrial Dysfunction in Autism Spectrum Disorder: S. Rose et al. Molecular diagnosis & therapy 2018; 22(5): 571–93.

43. Joseph RM, Fricker Z, Fenoglio A, Lindgren KA, Knaus TA, Tager-Flusberg H. Structural asymmetries of language-related gray and white matter and their relationship to language function in young children with ASD. Brain imaging and behavior 2014; 8(1): 60–72.

44. Li D, Karnath H-O, Xu X. Candidate biomarkers in children with autism spectrum disorder: a review of MRI studies. Neuroscience bulletin 2017; 33(2): 219–37.

45. Jones W, Klin A. Attention to eyes is present but in decline in 2–6-month-old infants later diagnosed with autism. Nature 2013; 504(7480): 427–31.

46. Mason L, Shic F, Falck-Ytter T, et al. Preference for biological motion is reduced in ASD: implications for clinical trials and the search for biomarkers. Molecular autism 2021; 12(1): 74.

47. Zamzow RM, Ferguson BJ, Ragsdale AS, Lewis ML, Beversdorf DQ. Effects of acute beta-adrenergic antagonism on verbal problem solving in autism spectrum disorder and exploration of treatment response markers. Journal of clinical and experimental neuropsychology 2017; 39(6): 596–606.

48. Curtin P, Austin C, Curtin A, et al. Dynamical features in fetal and postnatal zinc-copper metabolic cycles predict the emergence of autism spectrum disorder. Science advances 2018; 4(5): eaat1293.

49. Arora M, Reichenberg A, Willfors C, et al. Fetal and postnatal metal dysregulation in autism. Nature communications 2017; 8(1): 15493.

50. Jensen SS, Midya V, Arora M, et al. Pre-and postnatal trace element levels in primary teeth of children with and without an autism spectrum diagnosis. Environmental Research 2026; 295: 124001.

51. Ij H. Statistics versus machine learning. Nat Methods 2018; 15(4): 233.

52. Bae S, Hong J, Ha S, et al. Multimodal AI for risk stratification in autism spectrum disorder: integrating voice and screening tools. NPJ digital medicine 2025; 8(1): 538.

53. Austin C, Curtin P, Arora M, et al. Elemental dynamics in hair accurately predict future autism spectrum disorder diagnosis: an international multi-center study. Journal of Clinical Medicine 2022; 11(23): 7154.

54. Shmueli G. To explain or to predict? Statistical science 2010: 289–310.

55. Lai M-C, Lombardo MV, Auyeung B, Chakrabarti B, Baron-Cohen S. Sex/gender differences and autism: setting the scene for future research. Journal of the American Academy of Child & Adolescent Psychiatry 2015; 54(1): 11–24.

56. Corbett BA, Schwartzman JM, Libsack EJ, et al. Camouflaging in autism: Examining sex-based and compensatory models in social cognition and communication. Autism Research 2021; 14(1): 127–42.

57. Bölte S, Neufeld J, Marschik PB, Williams ZJ, Gallagher L, Lai M-C. Sex and gender in neurodevelopmental conditions. Nature Reviews Neurology 2023; 19(3): 136–59.

58. Posserud MB, Skretting Solberg B, Engeland A, Haavik J, Klungsøyr K. Male to female ratios in autism spectrum disorders by age, intellectual disability and attention-deficit/hyperactivity disorder. Acta Psychiatrica Scandinavica 2021; 144(6): 635–46.

59. Cheney AM, Barrera T, Rodriguez K, Jaramillo López AM. The intersection of workplace and environmental exposure on health in latinx farm working communities in rural Inland Southern California. International journal of environmental research and public health 2022; 19(19): 12940.

60. Hertz-Picciotto I, Croen LA, Hansen R, Jones CR, Van de Water J, Pessah IN. The CHARGE study: an epidemiologic investigation of genetic and environmental factors contributing to autism. Environmental health perspectives 2006; 114(7): 1119.

61. Hertz-Picciotto I, Schmidt RJ, Walker CK, et al. A prospective study of environmental exposures and early biomarkers in autism spectrum disorder: design, protocols, and preliminary data from the MARBLES study. Environmental health perspectives 2018; 126(11): 117004.

62. Berument SK, Rutter M, Lord C, Pickles A, Bailey A. Autism screening questionnaire: diagnostic validity. The British Journal of Psychiatry 1999; 175(5): 444–51.

63. Mullen EM. Mullen scales of early learning: AGS Circle Pines, MN; 1995.

64. Siper PM, Kolevzon A, Wang AT, Buxbaum JD, Tavassoli T. A clinician-administered observation and corresponding caregiver interview capturing DSM-5 sensory reactivity symptoms in children with ASD. Autism Research 2017; 10(6): 1133–40.

65. Colicino E, Cowell W, Pedretti NF, et al. Maternal steroids during pregnancy and their associations with exposure to lifetime stressful life events, prenatal stress appraisal and psychological functioning. Psychoneuroendocrinology 2023; 158: 106395.

66. Baron-Cohen S, Hoekstra RA, Knickmeyer R, Wheelwright S. The Autism-Spectrum Quotient (AQ)—Adolescent Version: Baron-Cohen, Hoekstra, Knickmeyer, and Wheelwright. Journal of autism and developmental disorders 2006; 36(3): 343–50.

67. Bölte S, Willfors C, Berggren S, et al. The roots of autism and ADHD twin study in Sweden (RATSS). Twin Research and Human Genetics 2014; 17(3): 164–76.

68. Ronald A, Larsson H, Anckarsäter H, Lichtenstein P. Symptoms of autism and ADHD: a Swedish twin study examining their overlap. Journal of abnormal psychology 2014; 123(2): 440.

69. Srivastava AK, Schwartz CE. Intellectual disability and autism spectrum disorders: causal genes and molecular mechanisms. Neuroscience & Biobehavioral Reviews 2014; 46: 161–74.

70. Michikawa T, Nitta H, Nakayama SF, et al. Baseline profile of participants in the Japan Environment and Children’s Study (JECS). Journal of Epidemiology 2018; 28(2): 99–104.

71. Rajabi P, Noori AS, Sargolzaei J. Autism spectrum disorder and various mechanisms behind it. Pharmacology Biochemistry and Behavior 2024; 245: 173887.

72. Mottron L, Bzdok D. Autism spectrum heterogeneity: fact or artifact? Molecular psychiatry 2020; 25(12): 3178–85.

73. Coco MI, Dale R. Cross-recurrence quantification analysis of categorical and continuous time series: an R package. Frontiers in psychology 2014; 5: 510.

74. Cofré R, Destexhe A. Entropy and complexity tools across scales in neuroscience: A review. Entropy 2025; 27(2): 115.

75. Schreiber T. Measuring information transfer. Physical review letters 2000; 85(2): 461.

76. Landa RJ, Reetzke R, Holingue CB, Herman D, Hess CR. Diagnostic stability and phenotypic differences among school-age children diagnosed with ASD before age 2. Frontiers in Psychiatry 2022; 13: 805686.

77. Ozonoff S, Young GS, Belding A, et al. The broader autism phenotype in infancy: when does it emerge? Journal of the American Academy of Child & Adolescent Psychiatry 2014; 53(4): 398–407. e2.

78. Wolpert DH. Stacked generalization. Neural networks 1992; 5(2): 241–59.

79. Geurts P, Ernst D, Wehenkel L. Extremely randomized trees. Machine learning 2006; 63(1): 3–42.

80. Lundberg SM, Lee S-I. A unified approach to interpreting model predictions. Advances in neural information processing systems 2017; 30.

81. Ozonoff S, Young GS, Carter A, et al. Recurrence risk for autism spectrum disorders: a Baby Siblings Research Consortium study. Pediatrics 2011; 128(3): e488–e95.

82. Constantino JN, Zhang Y, Frazier T, Abbacchi AM, Law P. Sibling recurrence and the genetic epidemiology of autism. American Journal of Psychiatry 2010; 167(11): 1349–56.

83. Akshoomoff N. Use of the Mullen Scales of Early Learning for the assessment of young children with autism spectrum disorders. Child Neuropsychology 2006; 12(4-5): 269–77.

84. McPheeters ML, Weitlauf A, Vehorn A, et al. Screening for autism spectrum disorder in young children: A systematic evidence review for the US Preventive Services Task Force. 2016.

85. Whalen S, Pandey G. A comparative analysis of ensemble classifiers: case studies in genomics. 2013 IEEE 13th International Conference on Data Mining; 2013: IEEE; 2013. p. 807–16.

