## Supplemental Material for "Machine-learning analysis of temporal molecular dynamics stratifies autism likelihood - a multinational study"

### **Supplementary Material**

| **Table of Contents** | **Page Number** |
| --- | --- |
| **Supplementary Methods and Additional Results** | 2-6 |
| ***Section S1***: Additional Details on Laboratory Protocols | 2-3 |
| ***Section S2:*** Additional Details on the Study Populations | 3-4 |
| ***Section S3:*** Feature Extraction Methodology | 4-6 |
| ***Section S4:*** Threshold Tuning and Statistical Analysis | 6 |
| **Supplementary Tables** | 7-12 |
| ***Table S1***: Cohort Breakdown showing distribution of sex, samples, age at hair collection, and Autism prevalence at different modeling stages | 7 |
| ***Table S2***: Full Stage-wise metrics for our likelihood stratification framework | 8 |
| ***Table S3***: Sex-stratified Stage-wise and Overall performance metrics when excluding JECS and PRISM (non-autism only cohorts) | 9 |
| ***Table S4:*** Sex-stratified Stage-wise and Overall performance metrics when excluding samples 21 years and older | 10 |
| ***Table S5:*** Sex-stratified Stage-wise and Overall performance metrics when excluding CHARGE and MARBLES samples with Mullen Scales Early Learning score < 70 | 11 |
| ***Table S6***: Sex-stratified Stage-wise and Overall performance metrics when performing bootstrap aggregation in the stacking layer. | 12 |
| **Supplementary Figures** | 13-24 |
| ***Figure S1***: Plots for Males and Females showing the performance on the validation set when we vary the probability thresholds in (A) stage 1 and (B) stage 2 within the range that met our set target criteria on their respective tuning sets. | 13 |
| ***Figure S2:*** Boxplot showing the distribution of Ca:S ratio across all cohorts and categorized by location | 14 |
| ***Figure S3:*** Boxplot showing the distribution of Cu:S ratio across all cohorts and categorized by location | 15 |
| ***Figure S4:*** Boxplot showing the distribution of Li:S ratio across all cohorts and categorized by location | 16 |
| ***Figure S5:*** Boxplot showing the distribution of Mg:S ratio across all cohorts and categorized by location | 17 |
| ***Figure S6:*** Boxplot showing the distribution of Mn:S ratio across all cohorts and categorized by location | 18 |
| ***Figure S7:*** Boxplot showing the distribution of P:S ratio across all cohorts and categorized by location | 19 |
| ***Figure S8:*** Boxplot showing the distribution of Sr:S ratio across all cohorts and categorized by location | 20 |
| ***Figure S9:*** Boxplot showing the distribution of Zn:S ratio across all cohorts and categorized by location | 21 |
| ***Figure S10:*** Boxplot showing the distribution of As:S ratio across all cohorts and categorized by location | 22 |
| ***Figure S11:*** Boxplot showing the distribution of Pb:S ratio across all cohorts and categorized by location | 23 |
| ***Figure S12:*** Boxplot showing the distribution of Ba:S ratio across all cohorts and categorized by location | 24 |

#### Supplementary Methods and Additional Results

##### Section S1: Additional Details on Laboratory Protocols

Elemental measurements were conducted at two laboratories (*LinusBio* Clinical Laboratory and Icahn School of Medicine at Mount Sinai) using comparable LA-ICP-MS workflows. Both labs employed similar hair preparation procedures, daily instrument tuning criteria, oxide and fractionation monitoring thresholds, and sulfur normalization of elemental time series. Laboratory-specific instrumentation parameters and quality control procedures are described below. Elemental signals from both laboratories were processed using identical element-to-sulfur normalization procedures, which helps mitigate variability arising from differences in hair density, ablation efficiency, or instrument sensitivity.

###### S1.A Linusbio Clinical Laboratory

**Hair Sample Preparation**

A single hair strand was selected from each participant and washed in a 1% Triton X-100 solution prepared with ultrapure water (18.2 MΩ·cm⁻¹) under ultrasonication for 1 minute. The strands were then rinsed several times with ultrapure water to remove residual surfactant, dried in an oven at 60 °C overnight, and mounted on plain glass microscope slides using double-sided tape. For longer strands, samples were trimmed from the distal end to fit on the microscope slide, typically to approximately 3 cm. In most cases, ablation was performed over a 1.2 cm segment beginning at the scalp/root end. For shorter strands (< 1.2 cm), the available length was used.

**LA-ICP-MS Analysis**

Elemental analysis of the hair strands was performed using a laser ablation (LA) system equipped with a 193 nm excimer laser (Elemental Scientific Lasers, Elemental Scientific (ESI)) coupled to an inductively coupled plasma-mass spectrometer (ICP-MS, Agilent Technologies, Inc.). Samples were ablated under a flow of helium, which was mixed with argon via a Y-piece prior to introduction into the ICP-MS.

**Instrument Calibration and Quality Control**

To ensure analytical accuracy, matrix-matched standards containing sulfur (S) levels equivalent to human hair were prepared from a mixture of poly(vinyl alcohol), glycerol, and K-carrageenan and doped with a mixture of metals at varying levels. In addition, three QC standards were prepared and spiked independently with the elements at low, medium, and high concentrations. These standard materials were used to verify that LA-ICP-MS instruments were within control whilst samples were being tested. The standards were produced at the Icahn School of Medicine at Mount Sinai, and concentrations were verified externally after acid digestion. The standards were characterized in the *LinusBio* clinical laboratory under the same method parameters as hair samples.

The National Institute of Standards and Technology certified reference material NIST612 Trace Elements in glass was used at the beginning of each day to tune the LA-ICP-MS. Daily sensitivity was optimized using 7, 55, 66, 88, 138, and 208 m/z, while ensuring oxide formation (^232^Th^16^O^+^/^232^Th^+^, < 0.3%) and fractionation (^232^Th^+^/^238^U^+^, 100 ± 6%) were within the specified limits. Instrument (LA-ICP-MS) performance was continuously assessed using the matrix-matched standards. Calibration curves were constructed using at least three standards for each element, and linearity and percent deviation from expected values were reviewed in *LinusBio*’s in-house proprietary software for implementing instrument calibration and statistical quality control processes. QC materials were run at least every 3 hours, and instrument performance was assessed before samples were cleared to run. Instrument performance was also monitored using control charts of the mean concentration of the QC materials, in accordance with *LinusBio*’s internal quality protocol. Data that passed QA/QC was cleared for downstream processing.

###### S1.B Icahn School of Medicine at Mount Sinai

Full details have been described previously.^1^ A summary of the key steps is provided below.

**Hair Sample Preparation**

Single hair strands from each participant were washed to remove surface contaminants in a solution of 1% Triton X-100 and ultrapure water (18.2 MΩ cm⁻¹) using sonication for 1 minute. Hairs were then rinsed with ultrapure water to remove the surfactant and dried in an oven at 60 °C overnight. Hairs were mounted on plain glass microscope slides using double-sided tape and loaded into an ablation cell.

**LA-ICP-MS Analysis**

Elemental analysis was performed using a New Wave Research NWR-193 laser ablation unit (ESI, Beaverton, OR, USA) equipped with a 193 nm ArF excimer laser, coupled to an Agilent 8800 triple-quadrupole ICP-MS (Agilent Technologies, Santa Clara, CA, USA). Helium was used as a carrier gas from the laser ablation cell and mixed with argon via a Y-piece before introduction into the ICP-MS. The system was tuned daily using NIST SRM 612 (Trace Elements in Glass) to monitor sensitivity (maximum analyte ion counts), oxide formation (²³²Th¹⁶O⁺/²³²Th⁺ < 0.3%), and fractionation (²³²Th⁺/²³⁸U⁺ = 100 ± 5%).

**Laser Ablation Procedure and Signal Processing**

A pre-ablation scan at low laser energy (0.27–0.32 J cm⁻²) was first run along the hair to remove the surface layer and reduce contamination of the endogenous signal. The hair was then scanned along the same path at higher energy (0.50–0.55 J cm⁻²) to collect elemental signal intensity along the strand. Approximately 10 mm of hair was scanned along each strand, representing about one month of growth and providing over 650 sampling points.

##### Section S2: Additional Details on the Study Populations

**CHARGE and MARBLES:** CHARGE and MARBLES study protocols were approved by the University of California at Davis and State of California Institutional Review Boards.

The CHARGE protocol collected a hair sample at the study visit and also asked the parent to provide a few strands of hair from the child’s first haircut, if they had saved that hair. Hair samples from both time points were collected for 38% of the CHARGE participants. Hair samples were evaluated for hair quality, and then a random selection if both were available, selecting the clinic-collected hair 64% of the time. In total, 26% of CHARGE samples were from the child’s first haircut. The MARBLES protocol attempted to collect hair samples at 3, 12, and 24 months, with samples from multiple time points sent for 92% of MARBLES participants. Again, hair samples were inspected for hair quality, and then the sample from the latest time point was typically selected (~95% of the time). In addition to assessing children for autism, those in both MARBLES and CHARGE were assessed for developmental delay using the Mullen Scales of Early Learning (MSEL), a standardized instrument for ages 3 to 60 months that assesses cognitive development.

**SEAVER and PRISM**: The present study was approved by the Mount Sinai IRB, and written informed consent was obtained from all participants and/or their parents or guardians.

**RATSS**: This study was approved by the Swedish Regional Ethical Review Board.

**JECS**: This protocol was reviewed and approved by the IRB of the Japanese Ministry of the Environment’s on Epidemiological Studies and by the Ethics Committees of all participating institutions. All participating women provided written informed consent.

**Mexico City**: This study was approved by the ethical review board of Iberoamericana University.

##### Section S3: Feature Extraction Methodology

***Time series preprocessing:*** Before feature extraction, elemental time series underwent a standardized preprocessing pipeline to enhance signal quality and comparability across samples. For each sample, the time series for each element was standardized and smoothed (optional for some features). Smoothing was carried out using a Savitzky-Golay filter with a window length of 25 time points and a polynomial order of 4. This is a signal processing technique that preserves local trends while reducing high-frequency noise^2^ Each time series was standardized to zero mean and unit variance, ensuring that downstream features reflected shape and dynamics rather than raw magnitude. Both smoothing and standardization were implemented in the Python library SciPy.^3^ Preprocessing was used selectively depending on the requirements of the feature extraction technique; for example, the *Summary Statistics* features were derived from raw, unprocessed time series.

The eight feature extraction techniques used in this analysis characterize various aspects of the temporality, including recurrence quantification analysis (RQA) and cross-recurrence quantification analysis (CRQA), that quantifies rhythmicity and synchronicity of elemental time series, entropy and complexity measures (Entropy-Complexity, Multiscale Cross-Entropy) that quantify irregularity and unpredictability in time series, causal network centrality (which assesses the structural importance of each element within a network inferred from inter-element causal relationships), distributional features (Summary Statistics), and methods that quantifies features of temporality (Catch22, TSFEL). Below, we provide detailed descriptions of each feature extraction technique.

***RQA (Recurrence Quantification Analysis):*** This is a nonlinear time series analysis technique used to quantify the temporal structure and recurrence behavior of dynamical systems. For each elemental time series, we computed a set of RQA features (13 in total) that capture dynamical characteristics such as recurrence rate, determinism, laminarity, etc.^4^ These measures were calculated over a range of recurrence radii to account for varying recurrence sensitivities. We then averaged each measure across the radii to obtain robust estimates for each measure, following the approach used in Panis 2023 et al.^5^ To construct the phase space required for RQA, the embedding dimension and time delay parameters were optimized for each time series using false nearest neighbors (FNN) and mutual information (MI), respectively, both standard approaches in nonlinear time series analysis.^6-8^ RQA computations were implemented using the Python library PyRQA, and the embedding dimension and time delay parameters were optimized using the NoLiTSA library.^9^

***CRQA (Cross Recurrence Quantification Analysis):*** CRQA extends RQA to pairs of time series, capturing dynamical coupling between them. For a pair of elements, CRQA quantifies the joint recurrence structures by analyzing a shared recurrence plot constructed from their embedded trajectories. As with RQA, we computed the same 13 recurrence measures across multiple recurrence radii and averaged them. And we optimized the embedding dimension and delay parameters using the same techniques. The same libraries were used.

***Entropy-Complexity:*** These features characterize the regularity (and unpredictability) and complexity of time-series data. For each element, we computed 5 features: its permutation entropy, spectral entropy, sample entropy, Hjorth mobility, and complexity using the Python package Antropy.^10^

***Multiscale Cross-Entropy:*** Multiscale cross-entropy (MSCE) characterizes the cross-entropy (i.e., the predictability of one signal from another) between two signals across multiple temporal scales. This was implemented using the EntropyHub library.^11^ Specifically, the *XMSEn* function was applied with the *cross-K2* entropy estimator.^11^ The number of scales was set to 3, resulting in 3 features per pair of elemental time series.

***Causal Network Centrality:*** This method quantifies the influence of individual elements in a multivariate system by measuring directed information flow between pairs of time series. Causal network centrality measures were derived from networks constructed using the PCMCI+ algorithm from the Python library Tigramite.^12^ In this context, each node in the network represents an element, and the edges represent the existence and strength of the causal relationship between them. Each network is converted into an adjacency structure, where 0 indicates no causal relationship between elements, and 1 indicates a causal relationship, regardless of strength, estimated temporal lags, or bidirectionality. Using the adjacency structure of this network, the following centrality measures (using the igraph library) were calculated for each element (node): betweenness centrality, eigenvector centrality, closeness centrality, degree centrality, and eccentricity.^13^

***Summary Statistics:*** Features generated here describe the distributional characteristics and dispersion of each elemental time series. For each elemental time series, we compute its first four moments: mean, standard deviation, skewness, and kurtosis. These metrics characterize central tendency, variability, asymmetry, and peakedness, respectively. In addition, we extract a set of percentile-based features to describe the distribution's spread and shape, including the interquartile range and quantiles at the 0th, 10th, 25th, 50th, 75th, 90th, and 100th percentiles. These metrics capture the range and skew of the distribution in a non-parametric way. This approach results in 12 features per time series.

***CATCH22 (CAnonical Time-series CHaracteristics):*** CATCH22 features are a set of 22 statistical properties of time series data from the pycatch22 Python library.^14^ These features capture a diverse range of statistical, temporal, and nonlinear properties. The features were selected through performance-based screening from a large pool of thousands and include time series properties such as distributional shape, autocorrelation structure, and periodicity. We used the pycatch22 library to compute 22 features for each element.

***TSFEL (Time Series Feature Extraction Library):*** TSFEL generates 162 features for a time series.^15^ These features are based on statistical, temporal, spectral, and fractal properties of time series data.

##### Section S4: Threshold Tuning and Statistical Analysis

Extra Details on stage 1: Since the probability distributions may differ between the tuning and validation sets, we fitted a polynomial function that maps raw predicted probabilities to a scaled version (ranging from 0 to 1), preserving the existing ranks within the data. Both the scaling and the polynomial function were learned using the pooled raw predicted probabilities across both male and female subjects in the tuning set. Analysis was conducted in Python (version 3.10). Initial data cleaning was performed using the *pandas* library.^16^ Modeling was performed with the *scikit-learn* library. ^17^

***Prevalence-adjusted predictive values***

Below are formulae used to compute prevalence-adjusted predictive values.

$${PPV}_{adjusted}= \frac{sensitivity \times prevalence}{(sensitivity \times prevalence) + ((1-specificity) \times(1-prevalence))}$$

$${NPV}_{adjusted}= \frac{specificity \times(1-prevalence)}{(specificity \times(1-prevalence)) + ((1-sensitivity) \times prevalence)}$$

#### Supplementary Tables

##### Table S1: Cohort Breakdown showing distribution of sex, samples, age at hair collection, and Autism prevalence at different modeling stages

| **Modelling Phase** | **Cohort (Country)** | **Sex** | **Sample size** | **Age at hair collection, *median (IQR)*** | **Autism Prevalence (%)** |
| --- | --- | --- | --- | --- | --- |
| **Training** | RATSS (Sweden) | Male | 76 | 180.0 (63.0) | 34.21% |
|  |  | Female | 62 | 168.0 (60.0) | 25.81% |
|  | Overall |  | 138 |  |  |
| **Stacking*** | CHARGE (CA, USA) | Male | 271 | 26.0 (18.0) | 42.80% |
|  |  | Female | 68 | 31.0 (9.0) | 25.00% |
|  | MARBLES (CA, USA) | Male | 128 | 12.54 (11.63) | 33.60% |
|  |  | Female | 104 | 12.49 (11.755) | 16.35% |
|  | Overall |  | 571 |  |  |
| **Tuning**** | CHARGE (CA, USA) | Male | 295 | 47.0 (11.5) | 0.00% |
|  |  | Female | 111 | 48.0 (10.5) | 0.00% |
|  | MARBLES (CA, USA) | Male | 1 | 37.04 (0.0) | 0.00% |
|  |  | Female | 1 | 36.08 (0.0) | 0.00% |
|  | RATSS (Sweden) | Male | 76 | 180.0 (63.0) | 34.21% |
|  |  | Female | 62 | 168.0 (60.0) | 25.81% |
|  | Overall |  | 546 |  |  |
| **Testing/**  **Validation** | CHARGE (CA, USA) | Male | 70 | 28.0 (21.0) | 44.29% |
|  |  | Female | 10 | 29.5 (7.25) | 0% |
|  | MARBLES (CA, USA) | Male | 31 | 12.88 (6.53) | 19.65% |
|  |  | Female | 22 | 12.24 (1.18) | 0% |
|  | RATSS (Sweden) | Male | 94 | 144.0 (84.0) | 24.47% |
|  |  | Female | 74 | 234. 0 (168.0) | 6.76% |
|  | Mexico | Male | 34 | 35.0 (25.75) | 85.29% |
|  |  | Female | 12 | 37.0 (14.75) | 33.33% |
|  | SEAVER (NY, USA) | Male | 25 | 84 (84.0) | 72% |
|  |  | Female | 14 | 72 (45.0) | 21.43% |
|  | PRISM (NY, USA) | Male | 51 | 48 (6.5) | 0% |
|  |  | Female | 33 | 47 (11.0) | 0% |
|  | JECS (Japan) | Male | 55 | 1 (0.0) | 0% |
|  |  | Female | 55 | 1 (0.0) | 0% |
|  | Overall |  | 580 |  |  |

*The stacking set was reused as the tuning set for the stage 2 model

**This refers to the tuning set used in stage 1 model

##### Table S2: Full Stage-wise metrics for our likelihood stratification framework^#^

| **Result on Stages of our Framework** | | | | | | | | | | |
| --- | --- | --- | --- | --- | --- | --- | --- | --- | --- | --- |
| **Stage 1: Low Likelihood Stratification** | | | | | | | | | | |
| **Sex** | **Sample Size**  **(% autism)** | **AUC***  **(95% CI)** | **Sensitivity**  *******  **(95% CI)** | **Specificity**  **(95% CI)** | **NPV at 14%****  **(95% CI)** | **NPV at 3%****  **(95% CI)** | **PPV at 14%****  **(95% CI)** | **PPV at 3%****  **(95% CI)** | **DLR+** | **DLR-** |
| Male | 360 (29.72) | 0.64  (0.57, 0.70) | 0.96  (0.91, 0.99) | 0.15  (0.11, 0.20) | 0.96  (0.86, 0.99) | 0.99  (0.92, 1.00) | 0.16 (0.12, 0.20) | 0.03 (0.02, 0.06) | 1.13 | 0.26 |
| Female | 220 (5.45) | 0.69  (0.56, 0.80) | 0.92  (0.65, 0.99) | 0.34  (0,28, 0.41) | 0.96  (0.89, 0.99) | 0.99  (0.94, 1.00) | 0.18 (0.13, 0.25) | 0.04 (0.02, 0.09) | 1.39 | 0.24 |
| Total | 580 (20.52) | 0.75  (0.70, 0.79) | 0.96  (0.91, 0.98) | 0.23  (0.20, 0.28) | 0.97  (0.92, 0.99) | 0.99  (0.96, 1.00) | 0.17 (0.14, 0.21) | 0.04 (0.02, 0.06) | 1.25 | 0.18 |
| **Stage 2: High and Intermediate Likelihood Stratification** | | | | | | | | | | |
| **Sex** | **Sample Size (% autism)** | **AUC***  **(95% CI)** | **Sensitivity**  **(95% CI)** | **Specificity*****  **(95% CI)** | **NPV at 14%****  **(95% CI)** | **NPV at 3%****  **(95% CI)** | **PPV at 14%** (95% CI)** | **PPV at 3%****  **(95% CI)** | **DLR+** | **DLR-** |
| Male | 319 (32.29) | 0.56 (0.49,0.62) | 0.23 (0.15, 0.31) | 0.86 (0.81,0.9) | 0.87 (0.86, 0.89) | 0.97 (0.97, 0.98) | 0.21 (0.14,0.3) | 0.05 (0.03,0.08) | 1.62 | 0.90 |
| Female | 148 (7.43) | 0.61 (0.46,0.76) | 0.09 (0.0, 0.3) | 0.96 (0.92,0.99) | 0.87 (0.85, 0.89) | 0.97 (0.97, 0.98) | 0.25 (0.0,0.65) | 0.06 (0.0,0.26) | 2.08 | 0.95 |
| Total | 467 (24.41) | 0.67 (0.62,0.72) | 0.22 (0.15, 0.29) | 0.9 (0.86,0.92) | 0.88 (0.87, 0.89) | 0.97 (0.97, 0.98) | 0.25 (0.17,0.34) | 0.06 (0.04,0.09) | 2.09 | 0.87 |

\# Note that although we present the sensitivity and specificity for each stage, reporting in such way essentially defeats the main benefits of a three-group stratification by collapsing them simply into two groups.

*Overall AUC is influenced by the high predicted probability of male autism cases and the low predicted probability of female non-autism samples. We estimated the overall AUC directly from predicted probabilities (rather than weighted averages) to facilitate thresholding for the rule-out test and because most cohorts are not sufficiently well-balanced between males and females to justify sex-specific weighting.

**NPV and PPV are reported at an autism prevalence level of 14% and 3%.

******* Since our framework is two-stage, the primary metrics for stages 1 and 2 are sensitivity and specificity, respectively.

##### Table S3: Sex-stratified Stage-wise and Overall performance metrics when excluding JECS and PRISM (non-autism only cohorts)*

| **Stage 1: Low Likelihood Stratification** | | | | | | |
| --- | --- | --- | --- | --- | --- | --- |
| **Sex** | **Sample Size** | **Num. Cases** | **AUC****  **(95% CI)** | **Sensitivity (95% CI)** | **NPV@14*****  **(95% CI)** | **NPV@3*****  **(95% CI)** |
| Male | 254 | 107 | 0.66  (0.59, 0.73) | 0.96  (0.91, 0.99) | 0.96  (0.85, 0.99) | 0.99 (0.90, 1.00) |
| Female | 132 | 12 | 0.73  (0.58, 0.84) | 0.92  (0.65, 0.99) | 0.97  (0.86, 0.99) | 0.99 (0.91, 1.00) |
| Overall | 386 | 119 | 0.76  (0.71, 0.81) | 0.96  (0.91, 0.98) | 0.97  (0.92, 0.99) | 0.99 (0.95, 1.00) |
| **Stage 2: High and Intermediate Likelihood Stratification** | | | | | | |
| **Sex** | **Sample Size** | **Num. Cases** | **AUC****  **(95% CI)** | **Specificity 95% CI)** | **PPV@14*****  **95% CI)** | **PPV@3*****  **( 95% CI)** |
| Male | 226 | 103 | 0.56  (0.48, 0.63) | 0.89  (0.83, 0.94) | 0.26 (0.14, 0.45) | 0.06 (0.02, 0.23) |
| Female | 86 | 11 | 0.58  (0.39, 0.74) | 0.93  (0.85, 0.97) | 0.18 (0.03, 0.58) | 0.04 (0.00, 0.45) |
| Overall | 312 | 114 | 0.66  (0.60, 0.72) | 0.91  (0.86, 0.94) | 0.28 (0.16, 0.45) | 0.07 (0.02, 0.22) |
| **Overall Performance** | | | | | | |
| **Sex** | **Sample Size** | **Num. Cases** | **Odds Ratio** | | | |
| Male | 254 | 107 | 3.08 (1.75,5.42) | | | |
| Female | 132 | 12 | 3.2 (0.99,10.31) | | | |
| Overall | 386 | 119 | 3.1 (1.86,5.17) | | | |

*PRISM (New York) and JECS (Japan) include all non-autism samples.

**Overall AUC is influenced by the high predicted probability of male autism cases and the low predicted probability of female non-autism samples. We estimated the overall AUC directly from predicted probabilities (rather than weighted averages) to facilitate thresholding for the rule-out test and because most cohorts are not sufficiently well-balanced between males and females to justify sex-specific weighting.

***NPV and PPV are reported at an autism prevalence level of 14% and 3%.

Since our framework is designed to be two-stage, the primary metrics for stages 1 and 2 are sensitivity and specificity, respectively.

##### Table S4: Sex-stratified Stage-wise and Overall performance metrics when excluding samples 21 years and older

| **Stage 1: Low likelihood Stratification** | | | | | | |
| --- | --- | --- | --- | --- | --- | --- |
| **Sex** | **Sample Size** | **Num. Cases** | **AUC***  **(95% CI)** | **Sensitivity (95% CI)** | **NPV@14****  **(95% CI)** | **NPV@3****  **(95% CI)** |
| Male | 343 | 105 | 0.65  (0.58, 0.71) | 0.96  (0.91, 0.99) | 0.96 (0.85, 0.99) | 0.99 (0.91, 1.00) |
| Female | 186 | 12 | 0.67  (0.55, 0.80) | 0.92  (0.65, 0.99) | 0.96 (0.88, 0.99) | 0.99 (0.93, 1.00) |
| Overall | 529 | 117 | 0.74  (0.69, 0.79) | 0.96  (0.90, 0.98) | 0.97 (0.92, 0.99) | 0.99 (0.96, 1.00) |
| **Stage 2: High and Intermediate Likelihood Stratification** | | | | | | |
| **Sex** | **Sample Size** | **Num. Cases** | **AUC***  **(95% CI)** | **Specificity 95% CI)** | **PPV@14****  **95% CI)** | **PPV@3****  **( 95% CI)** |
| Male | 305 | 101 | 0.55  (0.48, 0.62) | 0.85  (0.79, 0.89) | 0.20 (0.11, 0.33) | 0.05 (0.01, 0.15) |
| Female | 125 | 11 | 0.62  (0.44, 0.77) | 0.96  (0.91, 0.99) | 0.30 (0.07, 0.69) | 0.07 (0.01, 0.53) |
| Overall | 430 | 112 | 0.66  (0.60, 0.71) | 0.89  (0.85, 0.92) | 0.25 (0.15, 0.38) | 0.06 (0.02, 0.16) |
| **Overall Performance** | | | | | | |
| **Sex** | **Sample Size** | **Num. Cases** | **Odds Ratio** | | | |
| Male | 343 | 105 | 2.18 (1.37,3.48) | | | |
| Female | 186 | 12 | 4.0 (1.06,15.06) | | | |
| Overall | 529 | 117 | 2.34 (1.51,3.64) | | | |

*Overall AUC is influenced by the high predicted probability of male autism cases and the low predicted probability of female non-autism samples. We estimated the overall AUC directly from predicted probabilities (rather than weighted averages) to facilitate thresholding for the rule-out test and because most cohorts are not sufficiently well-balanced between males and females to justify sex-specific weighting.

**NPV and PPV are reported at an autism prevalence level of 14% and 3%.

Since our framework is designed to be two-stage, the primary metrics for stages 1 and 2 are sensitivity and specificity, respectively.

##### Table S5: Sex-stratified Stage-wise and Overall performance metrics when excluding CHARGE and MARBLES samples with Mullen Scales Early Learning score < 70***

| **Stage 1: Low Likelihood Stratification** | | | | | | |
| --- | --- | --- | --- | --- | --- | --- |
| **Sex** | **Sample Size** | **Num. Cases** | **AUC***  **(95% CI)** | **Sensitivity (95% CI)** | **NPV@14****  **(95% CI)** | **NPV@3****  **(95% CI)** |
| Male | 320 | 73 | 0.65 (0.57, 0.73) | 0.97 (0.91, 0.99) | 0.97 (0.87, 0.99) | 0.99 (0.91, 1.00) |
| Female | 216 | 12 | 0.69 (0.54, 0.79) | 0.92 (0.65, 0.99) | 0.96 (0.89, 0.99) | 0.99 (0.94, 1.00) |
| Overall | 536 | 85 | 0.74 (0.68, 0.79) | 0.96 (0.90, 0.99) | 0.98 (0.93, 0.99) | 1.00 (0.96, 1.00) |
| **Stage 2: High and Intermediate Likelihood Stratification** | | | | | | |
| **Sex** | **Sample Size** | **Num. Cases** | **AUC***  **(95% CI)** | **Specificity 95% CI)** | **PPV@14****  **(95% CI)** | **PPV@3****  **(95% CI)** |
| Male | 282 | 71 | 0.57 (0.50, 0.65) | 0.85 (0.80, 0.89) | 0.22 (0.12, 0.36) | 0.05 (0.01, 0.16) |
| Female | 145 | 11 | 0.61 (0.45, 0.75) | 0.96 (0.91, 0.98) | 0.25 (0.07, 0.60) | 0.06 (0.01, 0.43) |
| Overall | 427 | 82 | 0.66 (0.60, 0.73) | 0.89 (0.86, 0.92) | 0.26 (0.16, 0.39) | 0.06 (0.02, 0.17) |
| **Overall Performance** | | | | | | |
| **Sex** | **Sample Size** | **Num. Cases** | **Odds Ratio** | | | |
| Male | 320 | 73 | 2.49 (1.47,4.22) | | | |
| Female | 216 | 12 | 3.53 (1.02,12.19) | | | |
| Overall | 536 | 85 | 2.63 (1.62,4.28) | | | |

*Overall AUC is influenced by the high predicted probability of male autism cases and the low predicted probability of female non-autism samples. We estimated the overall AUC directly from predicted probabilities (rather than weighted averages) to facilitate thresholding for the rule-out test and because most cohorts are not sufficiently well-balanced between males and females to justify sex-specific weighting.

**NPV and PPV are reported at an autism prevalence level of 14% and 3%.

Since our framework is designed to be two-stage, the primary metrics for stages 1 and 2 are sensitivity and specificity, respectively.

***excluding intellectual disability and other psychiatric diagnoses (clinical diagnosis of intellectual disability was not available in this cohort; however, the Mullen early learning composite score was available for both CHARGE and MARBLES).

##### Table S6: Sex-stratified Stage-wise and Overall performance metrics when performing bootstrap aggregation in the stacking layer.

| **Stage 1: Low Likelihood Stratification** | | | | | | |
| --- | --- | --- | --- | --- | --- | --- |
| **Sex** | **Sample Size** | **Num. Cases** | **AUC* (95% CI)** | **Sensitivity**  **(95% CI)** | **NPV@14****  **(95% CI)** | **NPV@3****  **(95% CI)** |
| Male | 360 | 107 | 0.58 (0.50, 0.64) | 0.98 (0.93, 0.99) | 0.91 (0.62, 0.98) | 0.98 (0.72, 1.00) |
| Female | 220 | 12 | 0.63 (0.46, 0.77) | 0.92 (0.65, 0.99) | 0.94 (0.83, 0.98) | 0.99 (0.91, 1.00) |
| Overall | 580 | 119 | 0.72 (0.67, 0.77) | 0.97 (0.93, 0.99) | 0.97 (0.89, 0.99) | 0.99 (0.93, 1.00) |
| **Stage 2: High and Intermediate Likelihood Stratification** | | | | | | |
| **Sex** | **Sample Size** | **Num. Cases** | **AUC* (95% CI)** | **Specificity 95% CI)** | **PPV@14****  **( 95% CI)** | **PPV@3****  **( 95% CI)** |
| Male | 350 | 105 | 0.59 (0.52, 0.65) | 0.96 (0.92, 0.97) | 0.28 (0.13, 0.50) | 0.07 (0.01, 0.29) |
| Female | 173 | 11 | 0.61 (0.40, 0.77) | 1.00 (0.98, 1.00) | nan (nan, nan) | nan (nan, nan) |
| Overall | 523 | 116 | 0.70 (0.65, 0.75) | 0.97 (0.95, 0.98) | 0.36 (0.19, 0.58) | 0.10 (0.02, 0.34) |
| **Overall Performance** | | | | | | |
| **Sex** | **Sample Size** | **Num. Cases** | **Odds Ratio** | | | |
| Male | 360 | 107 | 2.25 (1.05,4.85) | | | |
| Female | 220 | 12 | 3.12 (0.39,24.83) | | | |
| Overall | 580 | 119 | 2.35 (1.16,4.79) | | | |

*Overall AUC is influenced by the high predicted probability of male autism cases and the low predicted probability of female non-autism samples. We estimated the overall AUC directly from predicted probabilities (rather than weighted averages) to facilitate thresholding for the rule-out test and because most cohorts are not sufficiently well-balanced between males and females to justify sex-specific weighting.

**NPV and PPV are reported at an autism prevalence level of 14% and 3%.

Since our framework is designed to be two-stage, the primary metrics for stages 1 and 2 are sensitivity and specificity, respectively.

#### Supplementary Figures

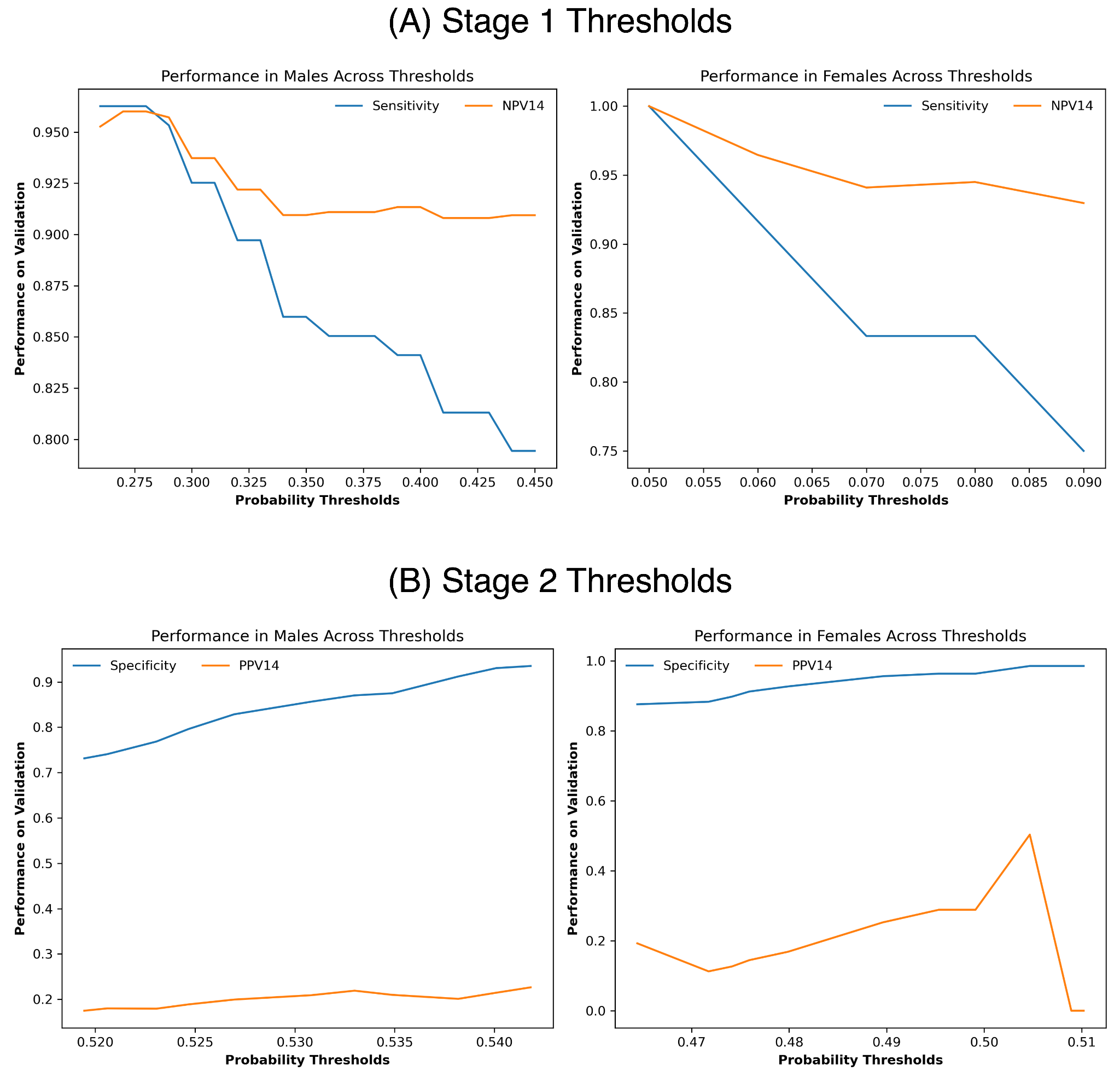

##### Figure S1: Plots for Males and Females showing the performance on the validation set when we vary the probability thresholds in (A) stage 1 and (B) stage 2 within the range that met our set target criteria on their respective tuning sets.

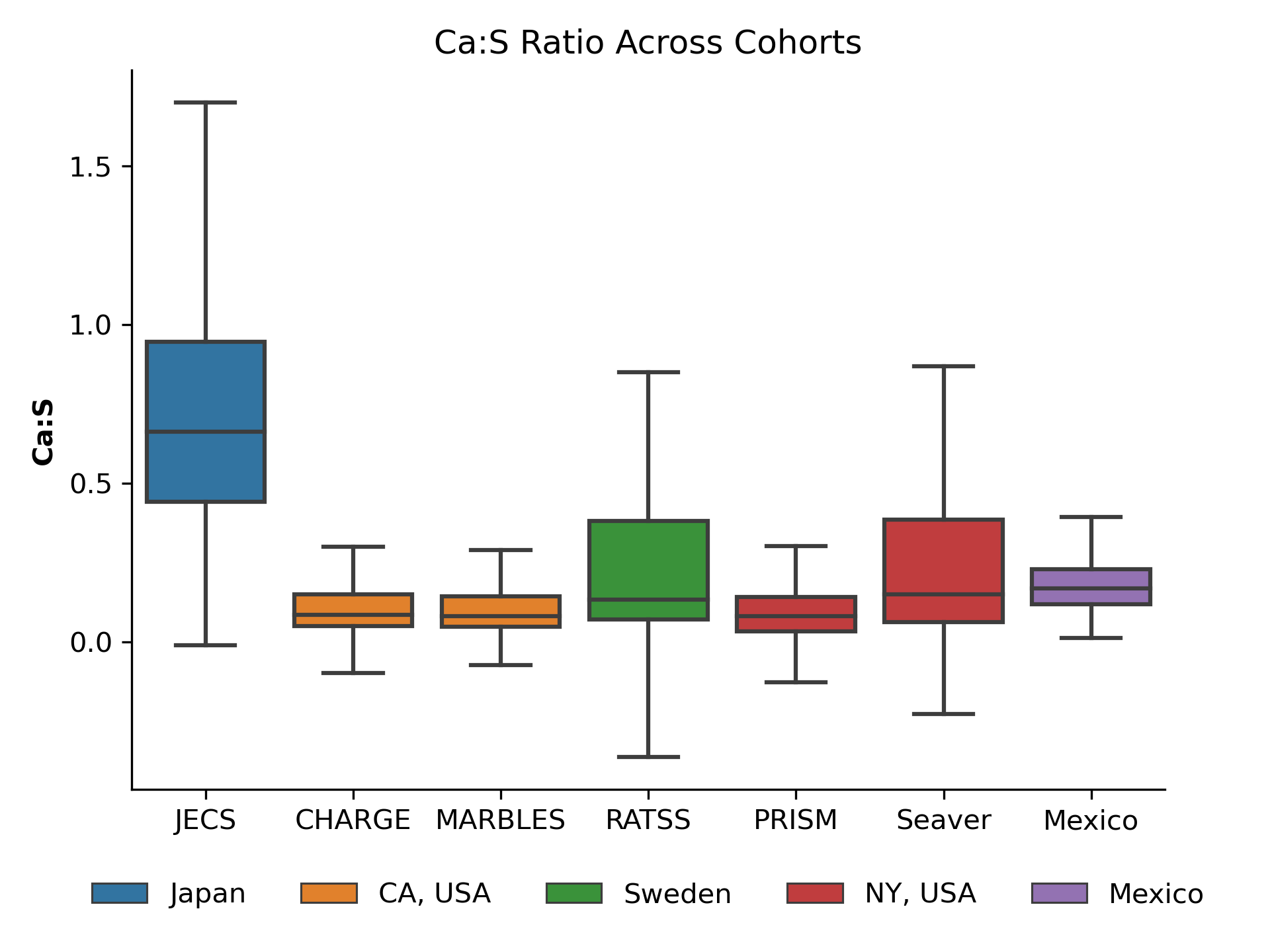

##### Figure S2: Boxplot showing the distribution of Ca:S ratio across all cohorts and categorized by location. Ca: Calcium, S: Sulfur, JECS: Japan Environment and Children’s Study, CHARGE: Childhood Autism Risks from Genetics and Environment Study, MARBLES: Markers of Autism Risk in Babies—Learning Early Signs Study, RATSS: Roots of Autism and ADHD Twin Study, PRISM: PRogramming of Intergenerational Stress Mechanisms Study.

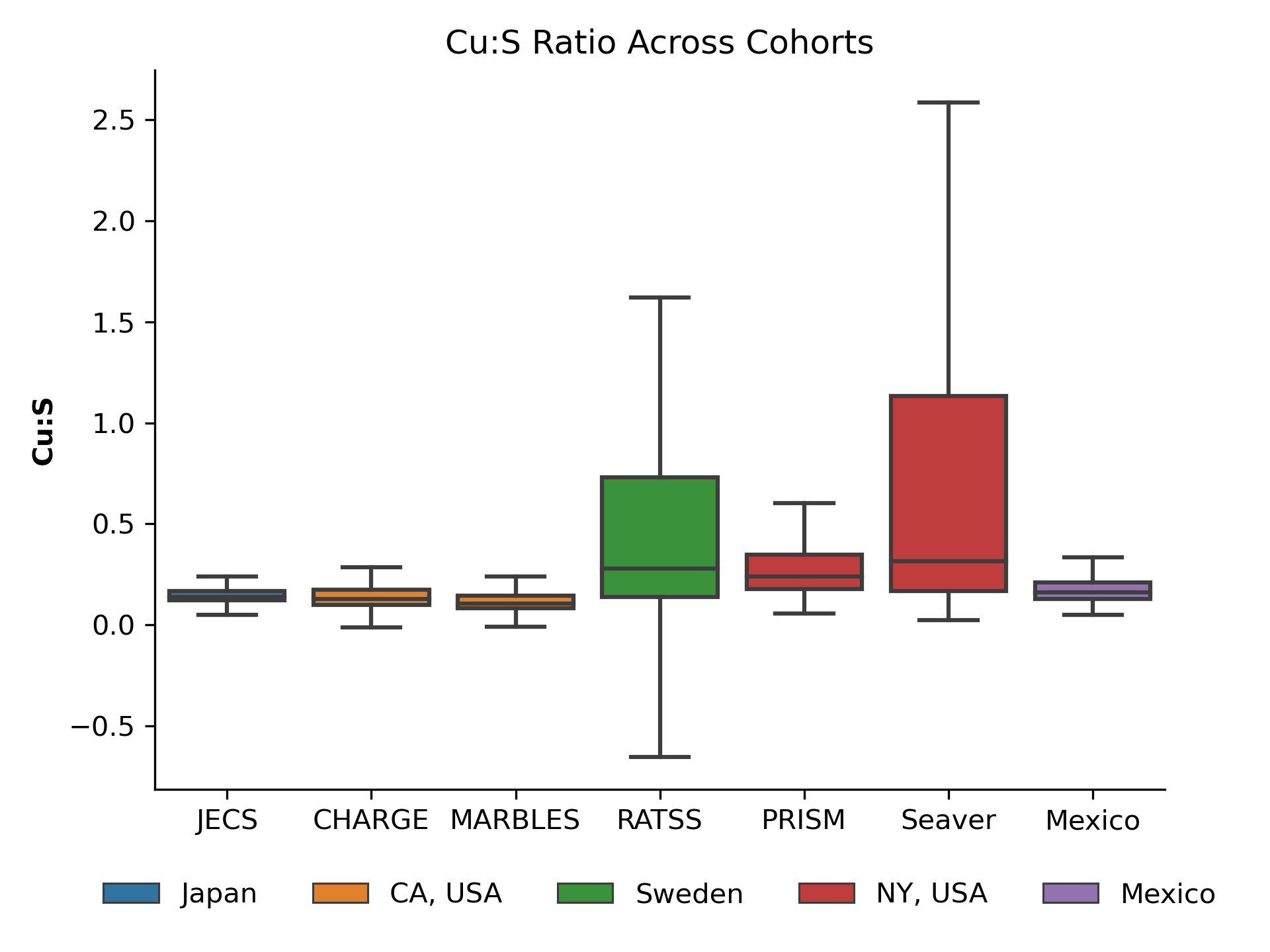

##### Figure S3: Boxplot showing the distribution of Cu:S ratio across all cohorts and categorized by location. Cu: Copper, S: Sulfur, JECS: Japan Environment and Children’s Study, CHARGE: Childhood Autism Risks from Genetics and Environment Study, MARBLES: Markers of Autism Risk in Babies—Learning Early Signs Study, RATSS: Roots of Autism and ADHD Twin Study, PRISM: PRogramming of Intergenerational Stress Mechanisms Study.

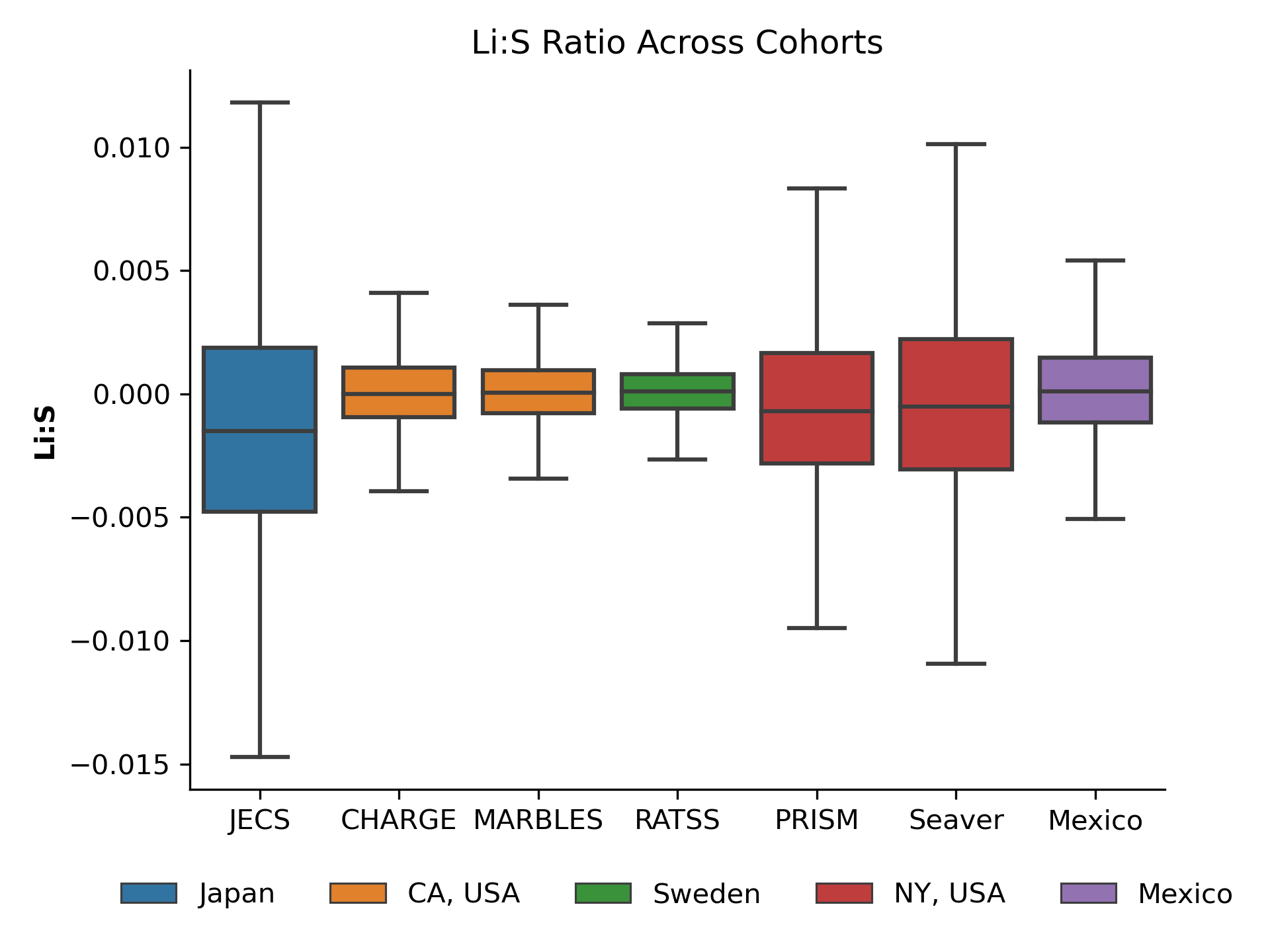

##### Figure S4: Boxplot showing the distribution of Li:S ratio across all cohorts and categorized by location. Li: Lithium, S: Sulfur, JECS: Japan Environment and Children’s Study, CHARGE: Childhood Autism Risks from Genetics and Environment Study, MARBLES: Markers of Autism Risk in Babies—Learning Early Signs Study, RATSS: Roots of Autism and ADHD Twin Study, PRISM: PRogramming of Intergenerational Stress Mechanisms Study.

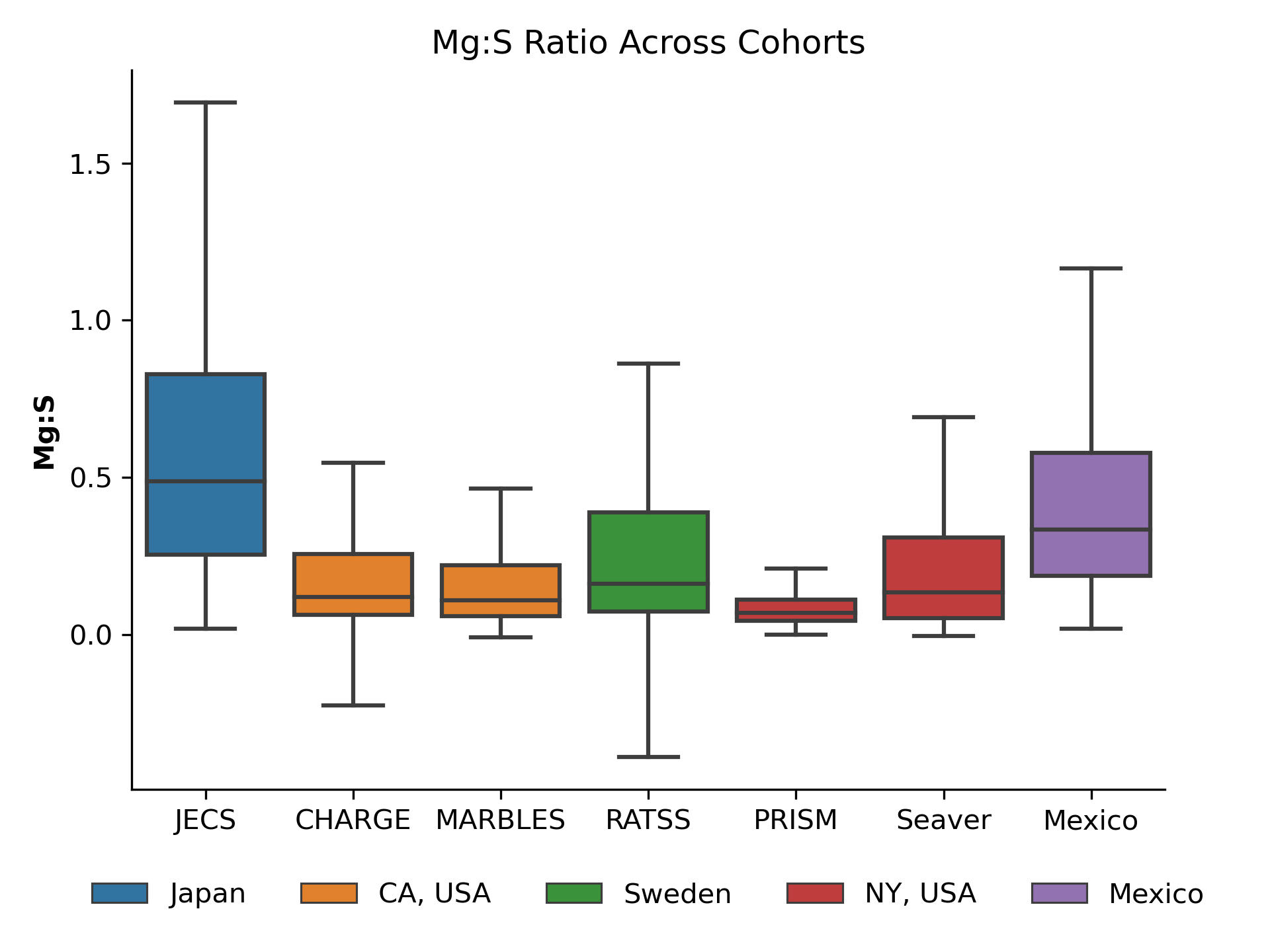

##### Figure S5: Boxplot showing the distribution of Mg:S ratio across all cohorts and categorized by location. Mg: Magnesium, S: Sulfur, JECS: Japan Environment and Children’s Study, CHARGE: Childhood Autism Risks from Genetics and Environment Study, MARBLES: Markers of Autism Risk in Babies—Learning Early Signs Study, RATSS: Roots of Autism and ADHD Twin Study, PRISM: PRogramming of Intergenerational Stress Mechanisms Study.

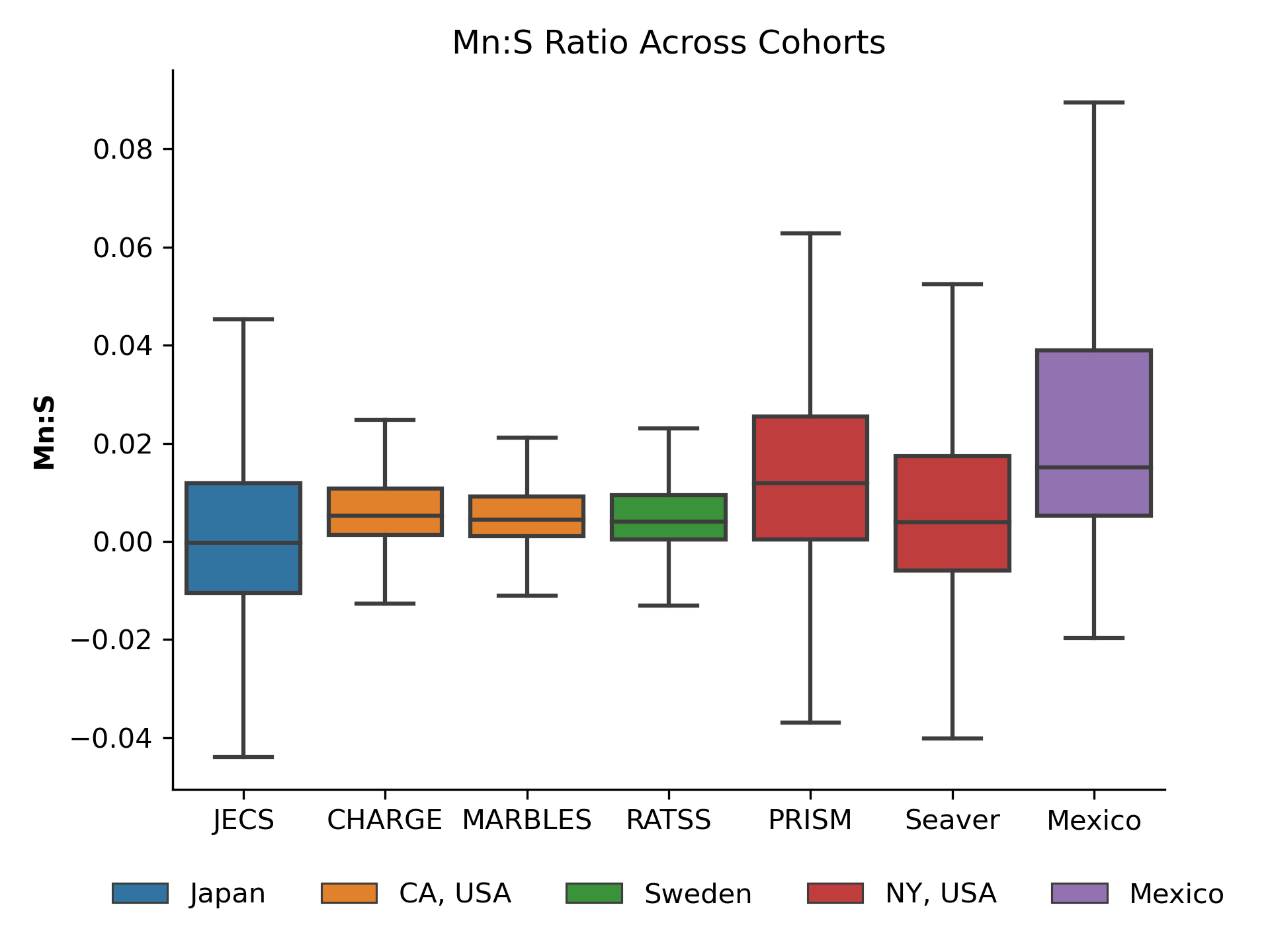

##### Figure S6: Boxplot showing the distribution of Mn:S ratio across all cohorts and categorized by location. Mn: Manganese, S: Sulfur, JECS: Japan Environment and Children’s Study, CHARGE: Childhood Autism Risks from Genetics and Environment Study, MARBLES: Markers of Autism Risk in Babies—Learning Early Signs Study, RATSS: Roots of Autism and ADHD Twin Study, PRISM: PRogramming of Intergenerational Stress Mechanisms Study.

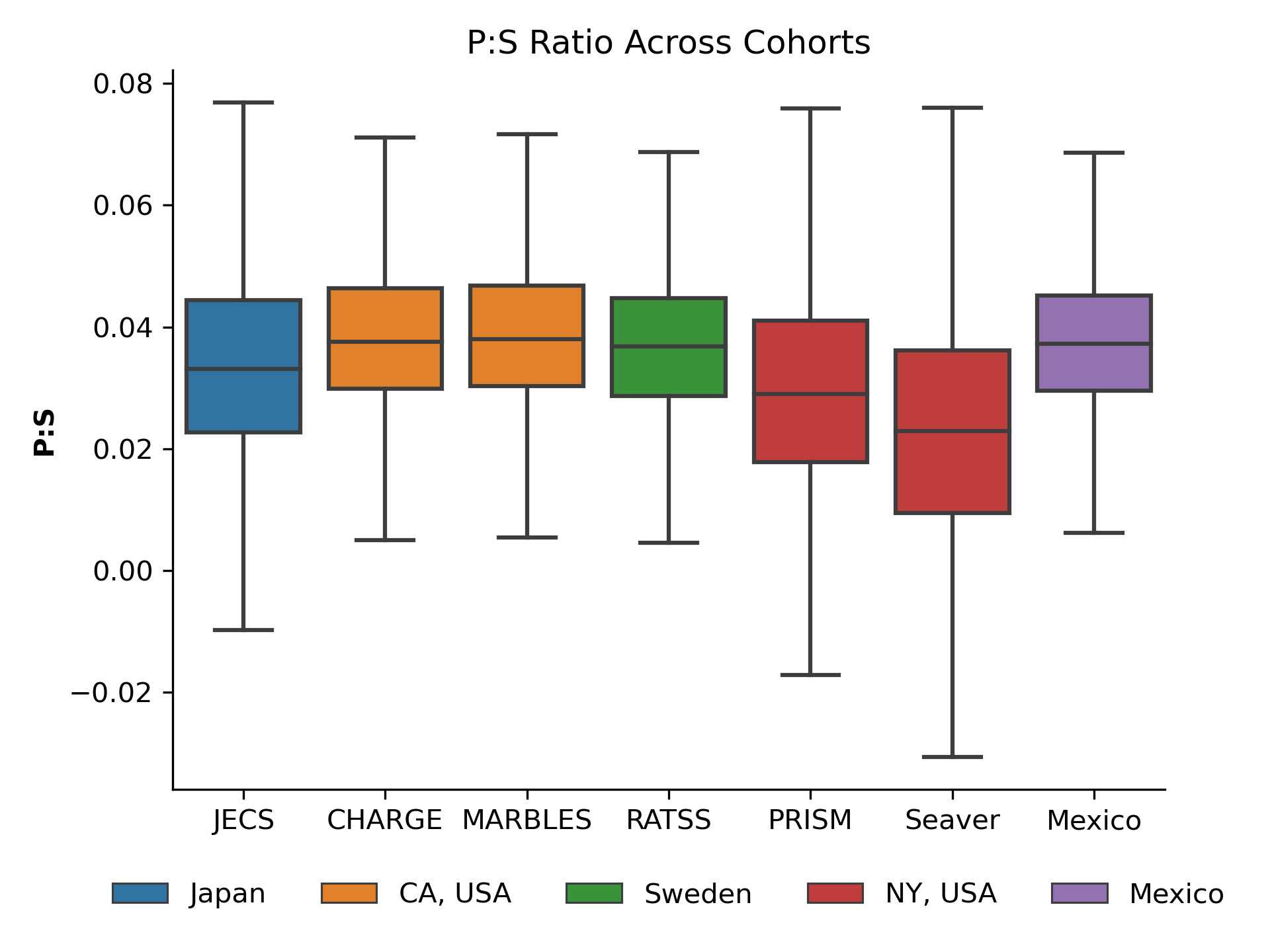

##### Figure S7: Boxplot showing the distribution of P:S ratio across all cohorts and categorized by location. P: Phosphorus, S: Sulfur, JECS: Japan Environment and Children’s Study, CHARGE: Childhood Autism Risks from Genetics and Environment Study, MARBLES: Markers of Autism Risk in Babies—Learning Early Signs Study, RATSS: Roots of Autism and ADHD Twin Study, PRISM: PRogramming of Intergenerational Stress Mechanisms Study.

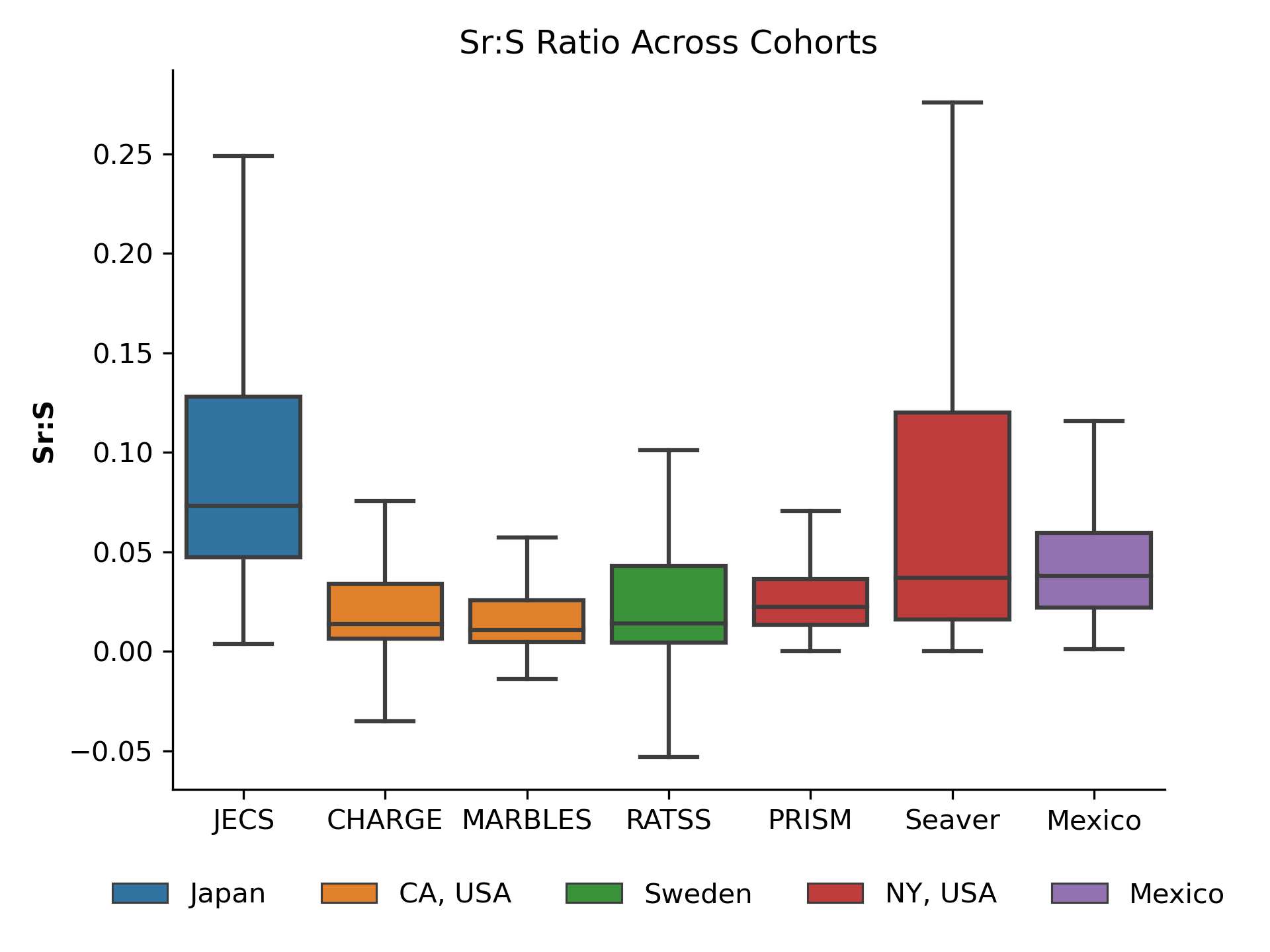

##### Figure S8: Boxplot showing the distribution of Sr:S ratio across all cohorts and categorized by location. Sr: Strontium, S: Sulfur, JECS: Japan Environment and Children’s Study, CHARGE: Childhood Autism Risks from Genetics and Environment Study, MARBLES: Markers of Autism Risk in Babies—Learning Early Signs Study, RATSS: Roots of Autism and ADHD Twin Study, PRISM: PRogramming of Intergenerational Stress Mechanisms Study.

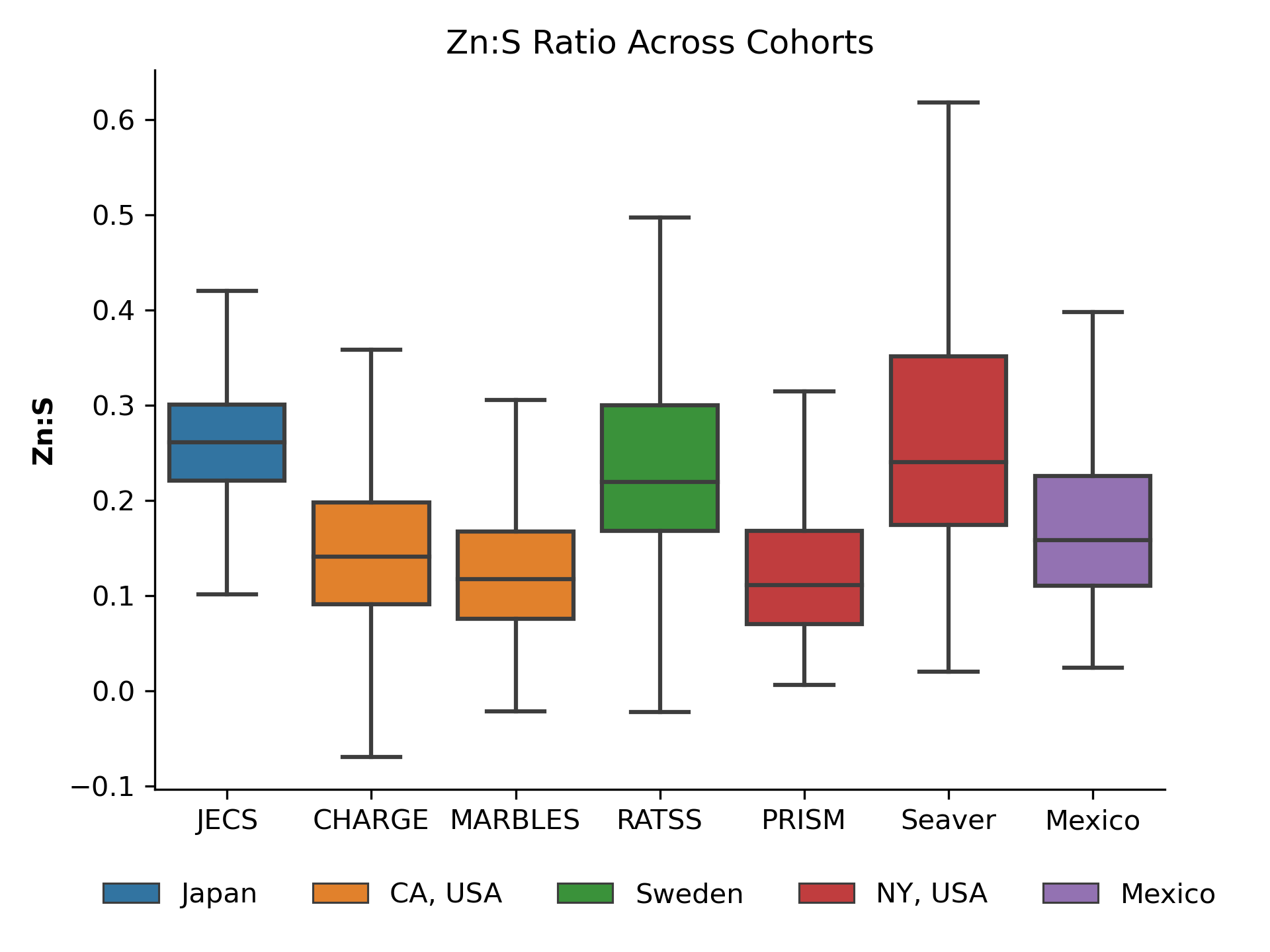

##### Figure S9: Boxplot showing the distribution of Zn:S ratio across all cohorts and categorized by location. Zn: Zinc, S: Sulfur, JECS: Japan Environment and Children’s Study, CHARGE: Childhood Autism Risks from Genetics and Environment Study, MARBLES: Markers of Autism Risk in Babies—Learning Early Signs Study, RATSS: Roots of Autism and ADHD Twin Study, PRISM: PRogramming of Intergenerational Stress Mechanisms Study.

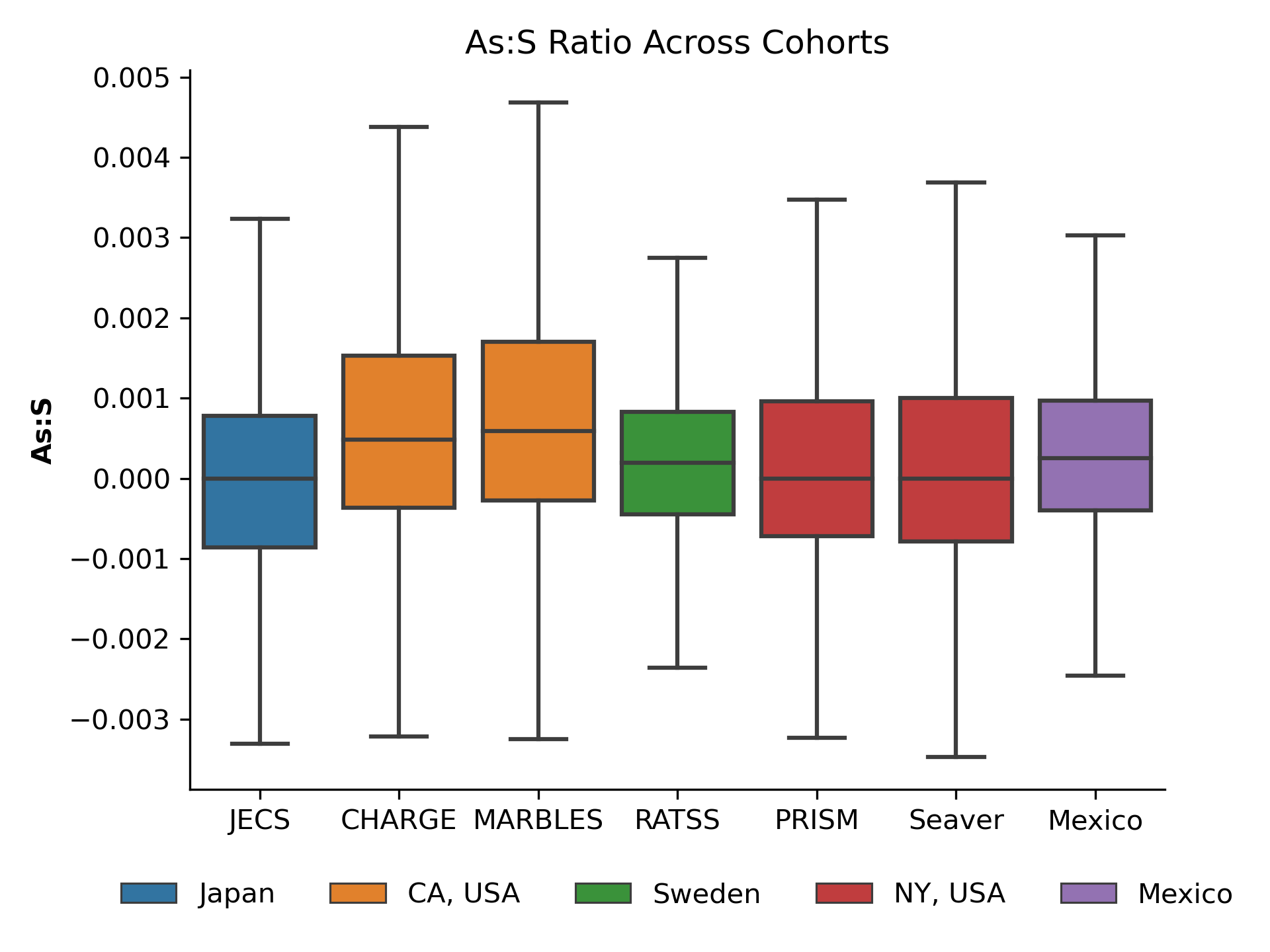

##### Figure S10: Boxplot showing the distribution of As:S ratio across all cohorts and categorized by location. As: Arsenic, S: Sulfur, JECS: Japan Environment and Children’s Study, CHARGE: Childhood Autism Risks from Genetics and Environment Study, MARBLES: Markers of Autism Risk in Babies—Learning Early Signs Study, RATSS: Roots of Autism and ADHD Twin Study, PRISM: PRogramming of Intergenerational Stress Mechanisms Study.

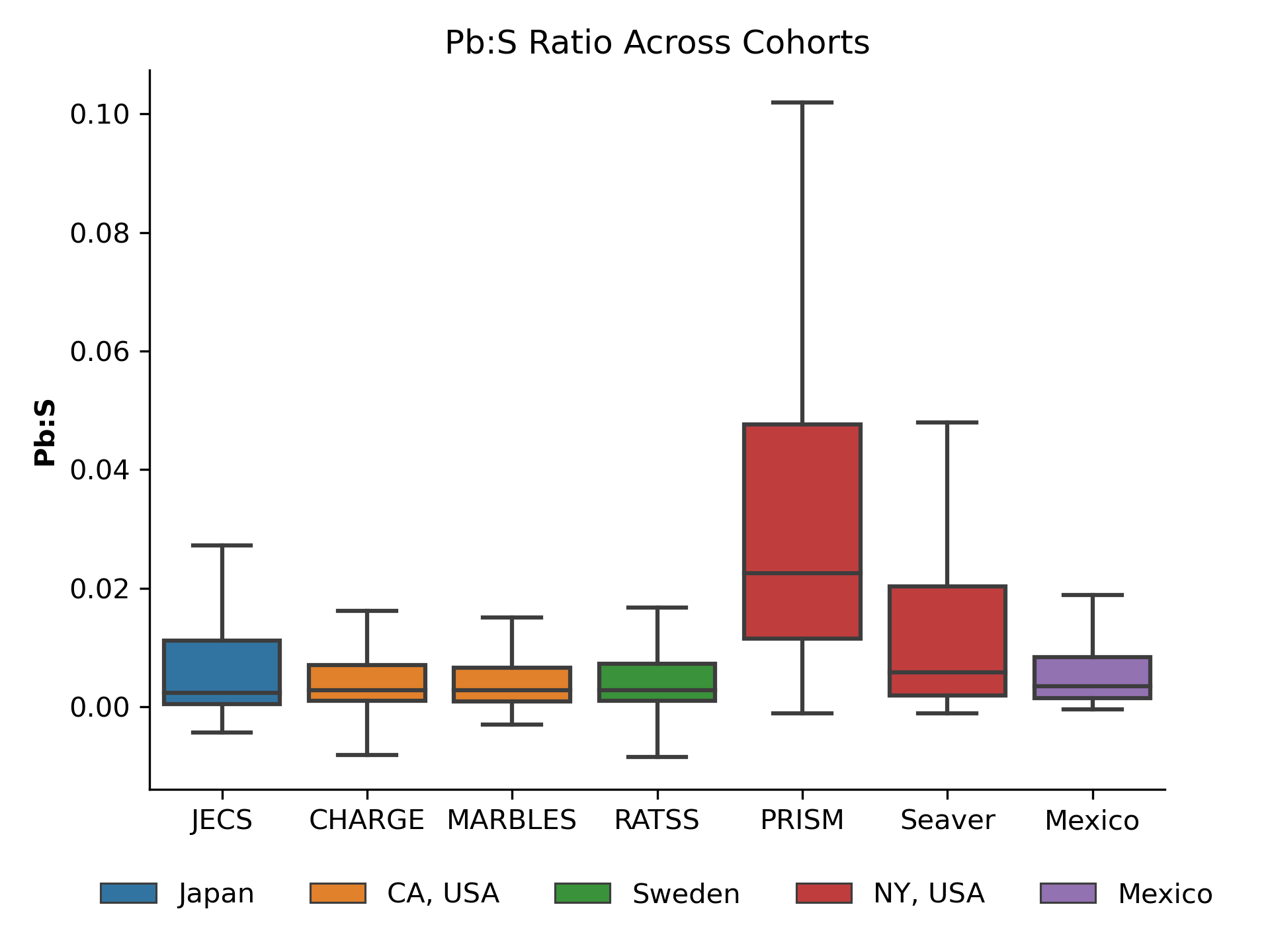

##### Figure S11: Boxplot showing the distribution of Pb:S ratio across all cohorts and categorized by location. Pb: Lead, S: Sulfur, JECS: Japan Environment and Children’s Study, CHARGE: Childhood Autism Risks from Genetics and Environment Study, MARBLES: Markers of Autism Risk in Babies—Learning Early Signs Study, RATSS: Roots of Autism and ADHD Twin Study, PRISM: PRogramming of Intergenerational Stress Mechanisms Study.

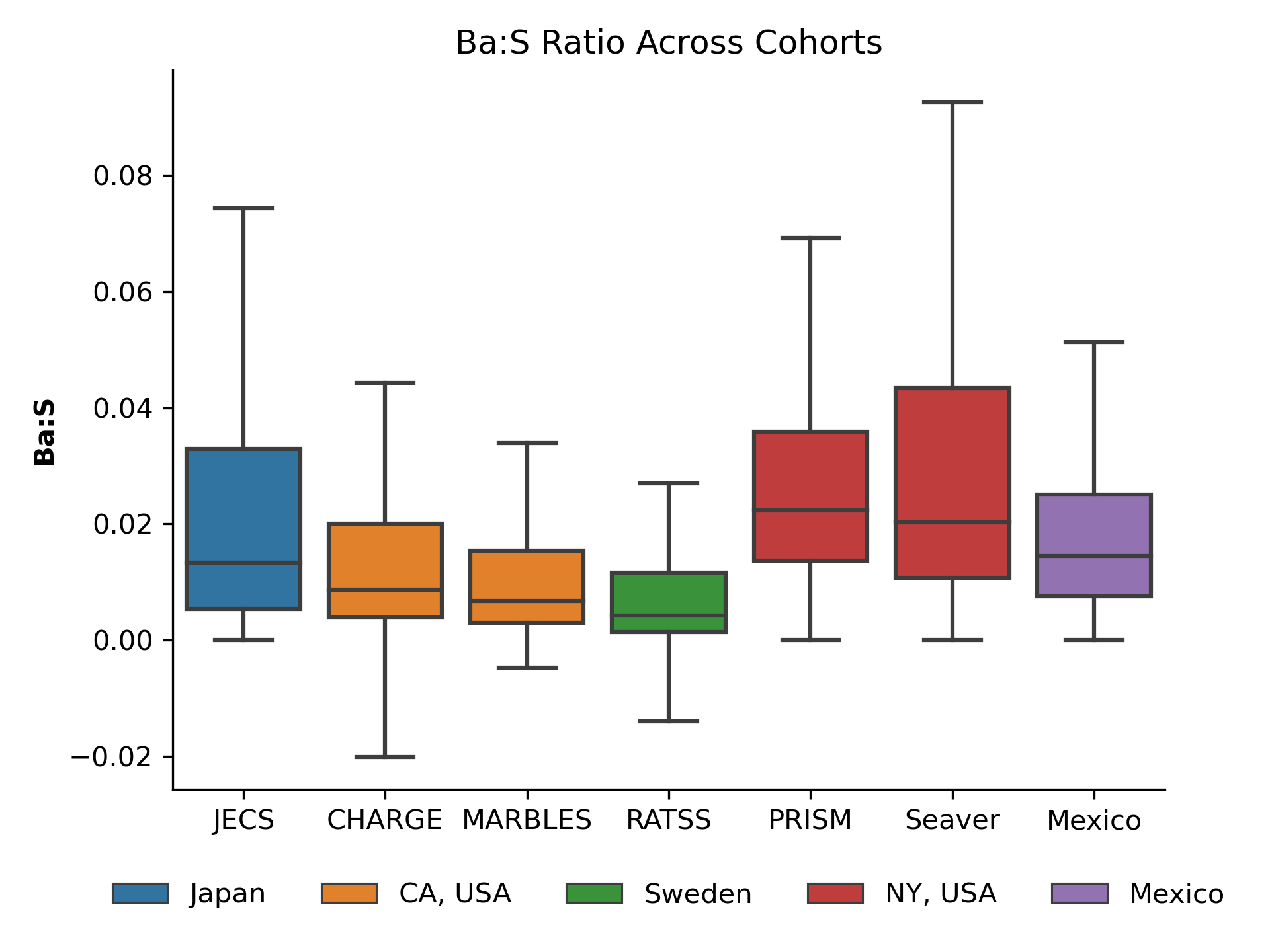

##### Figure S12: Boxplot showing the distribution of Ba:S ratio across all cohorts and categorized by location. Ba: Barium, S: Sulfur, JECS: Japan Environment and Children’s Study, CHARGE: Childhood Autism Risks from Genetics and Environment Study, MARBLES: Markers of Autism Risk in Babies—Learning Early Signs Study, RATSS: Roots of Autism and ADHD Twin Study, PRISM: PRogramming of Intergenerational Stress Mechanisms Study.

**References**

1.	Austin C, Curtin P, Arora M, et al. Elemental dynamics in hair accurately predict future autism spectrum disorder diagnosis: an international multi-center study. *Journal of Clinical Medicine* 2022; **11**(23): 7154.

2.	Savitzky A, Golay MJ. Smoothing and differentiation of data by simplified least squares procedures. *Analytical chemistry* 1964; **36**(8): 1627–39.

3.	Virtanen P, Gommers R, Oliphant TE, et al. SciPy 1.0: fundamental algorithms for scientific computing in Python. *Nature methods* 2020; **17**(3): 261–72.

4.	Marwan N. A historical review of recurrence plots. *The European Physical Journal Special Topics* 2008; **164**(1): 3–12.

5.	Pánis R, Adámek K, Marwan N. Averaged recurrence quantification analysis: Method omitting the recurrence threshold choice. *The European Physical Journal Special Topics* 2023; **232**(1): 47–56.

6.	Marwan N, Romano MC, Thiel M, Kurths J. Recurrence plots for the analysis of complex systems. *Physics reports* 2007; **438**(5-6): 237–329.

7.	Kennel MB, Brown R, Abarbanel HD. Determining embedding dimension for phase-space reconstruction using a geometrical construction. *Physical review A* 1992; **45**(6): 3403.

8.	Fraser AM, Swinney HL. Independent coordinates for strange attractors from mutual information. *Physical review A* 1986; **33**(2): 1134.

9.	Mannattil M, Pandey A, Verma MK, Chakraborty S. On the applicability of low-dimensional models for convective flow reversals at extreme Prandtl numbers. *The European Physical Journal B* 2017; **90**(12): 259.

10.	Vallat R. Antropy: Entropy and complexity of (EEG) time-series in Python. *GitHub repository* 2022.

11.	Flood MW, Grimm B. EntropyHub: An open-source toolkit for entropic time series analysis. *PloS one* 2021; **16**(11): e0259448.

12.	Runge J. Discovering contemporaneous and lagged causal relations in autocorrelated nonlinear time series datasets. Conference on uncertainty in artificial intelligence; 2020: Pmlr; 2020. p. 1388–97.

13.	Csardi G, Nepusz T. The igraph software. *Complex syst* 2006; **1695**: 1–9.

14.	Lubba CH, Sethi SS, Knaute P, Schultz SR, Fulcher BD, Jones NS. catch22: CAnonical Time-series CHaracteristics: Selected through highly comparative time-series analysis. *Data mining and knowledge discovery* 2019; **33**(6): 1821–52.

15.	Barandas M, Folgado D, Fernandes L, et al. TSFEL: Time series feature extraction library. *SoftwareX* 2020; **11**: 100456.

16.	McKinney W. Data structures for statistical computing in Python. *scipy* 2010; **445**(1): 51–6.

17.	Pedregosa F, Varoquaux G, Gramfort A, et al. Scikit-learn: Machine learning in Python.
